# Multi-Omics integration analysis of respiratory specimen characterizes baseline molecular determinants associated with COVID-19 diagnosis

**DOI:** 10.1101/2020.07.06.20147082

**Authors:** Jaswinder Singh Maras, Shvetank Sharma, Adil Bhat, Reshu aggrawal, Ekta Gupta, Shiv K Sarin

## Abstract

Rapid diagnosis and precise prognostication of SARS-CoV-2 infection remains a major challenge. A multi-omic approach was adopted, and in the discovery phase, global proteome/metaproteome/metabolome were analysed in the respiratory specimens of SARS-CoV-2 positive [n=20], negative [n=20], and H1N1 positive [n=5] cases. We identified MX1 (MX Dynamin Like GTPase 1) and WARS (Tryptophan--tRNA ligase) as clues to viral diagnosis and validated in 200 SARS-CoV-2 suspects. MX1 >30pg/ml and WARS >25ng/ml segregated virus positives patients [(AUC=94%CI(0.91-0.97)]. Distinct increase in SARS-CoV-2 induced immune activation, metabolic reprograming and a decrease in oxygen transport, wound healing, fluid regulation, vitamin and steroid metabolism was seen (p<0.05). Multi-omics profiling correlated with viraemia and segregated asymptomatic COVID-19 patients. Additionally, the multiomics approach identified increased respiratory pathogens [Burkholderiales, Klebsiella pneumonia] and decreased lactobacillus salivarius (FDR<0.05, p<0.05) in COVID-19 specimens.

**Conclusion:** Novel proteins [MX1 and WARS] can rapidly and reliably diagnose SARS-CoV-2 infection and identify asymptomatic and mild disease.

## Introduction

Severe acute respiratory syndrome coronavirus 2 (SARS-CoV-2), causes coronavirus disease 2019 (COVID-19). Spread of the virus is severe and within few months more than 5.69 millions were infected and mortality was seen in more than 356 thousand patient and counting. The global recovery rate of COVID-19 is nearly 41 % and the mortality rate is close to 7% (Ghinai et al., 2020; Guan et al., 2020; Li et al., 2007). About 80% of the patients infected with SARS-CoV-2 are asymptomatic or display mild symptoms and has good prognosis (Thevarajan et al., 2020) These patients may or may not require conventional medical treatment (Mehta et al., 2020; Thevarajan et al., 2020). However about 20% of patients who suffer from respiratory distress require immediate access to specialized intensive care; otherwise, they may die rapidly (Mehta et al., 2020; Murthy et al., 2020; Thevarajan et al., 2020; Wu and McGoogan, 2020). Further, limited understanding of SARS-CoV-2 pathogenesis delays speculative therapy for SARS-CoV-2 infection. Therefore, it is vital to understand the pathogenesis of SARS-CoV-2 infection and identify patients who are positive, asymptomatic or have very low viraemia as these patients are silent carrier of infection and could mediate an exponential increase in the infection rate. Further it is also important to stratify patients who are predisposed to COVID-19 associated severe respiratory distress. Therefore, employment of new approaches which could characterize baseline molecular determinants associated with SARS-CoV-2 diagnosis and prognosis becomes critical.

Currently oropharyngeal/nasopharyngeal swab (respiratory specimen) is used for RT-PCR based detection of viral genetic material and is the gold standard of COVID-19 detection; clinical sensitivity ranging from 66% to 80% (Udugama et al., 2020). However, the respiratory specimen is a mixture of viral, host molecules (proteins, metabolites) and microbiome which could be indicative of viral presence and could be utilized as a surrogate for screening purpose. Additionally, the respiratory specimen could work as a liquid biopsy for the diagnosis but also to give an insight into the pathophysiology of the COVID-19 disease. Methods with higher sensitivity and resolution than RT-PCR are needed to improve the management of the pandemic. Recently, Shen et.al (Shen et al., 2020) used high resolution mass spectrometry based proteomics and metabolomics in serum specimens to provide insights into the pathophysiology of COVID-19. However, detailed analysis and characterization of the respiratory specimen in terms of proteomics, metabolomics and metaproteomics needs to be analyzed for improving viral diagnostics and host related biomarkers of disease diagnosis and prognosis.

In this study, we hypothesized that SARS-CoV-2 induces characteristic molecular changes that can be detected in the respiratory specimen of COVID-19 positive patients. These molecular changes could be distinct and help segregate SARS-CoV-2 positive patients and provide pathophysiological insights which could be used for the development of new therapies. To test this hypothesis, global proteome, metaproteome and metabolome analysis was performed on the respiratory specimens followed by an in-depth analysis of the regulatory networks. The integrated analysis identified correlation clusters predictive of viral pathogenesis and promising targets for COVID-19 diagnosis. Targets identified in our derivative cohort, were validated in 200 COVID-19 suspected specimens and the sensitivity/specificity was recorded for COVID-19 diagnosis. We also analyzed the association of combined oropharyngeal and nasopharyngeal microbiome (metaproteome) with viral presence and SARS-CoV-2 diagnosis.

## Results

### Quantitation of SARS-CoV-2 and associated proteins in the respiratory specimen

#### Identification of viral diagnostic panel

We aimed to identify bio-molecules having diagnostic and prognostic value for SARS-CoV-2 infection (Figure 1A). Label free quantitative proteomics was performed and principle component analysis (PCA) followed by unsupervised clustering analysis showed clear distinction between the study groups (Figure 1B, Supplementary Figure 1, and Supplementary Table 1). Quantitative proteomics identified significant increase in 6 SARS-CoV-2 proteins along with ACE2 in the respiratory specimen of COVID-19 positive patients compared to negative patients (p<0.05, Figure 1C, H1N1 samples did not enrich any COVID-19 or associated proteins).

**Figure 1:**
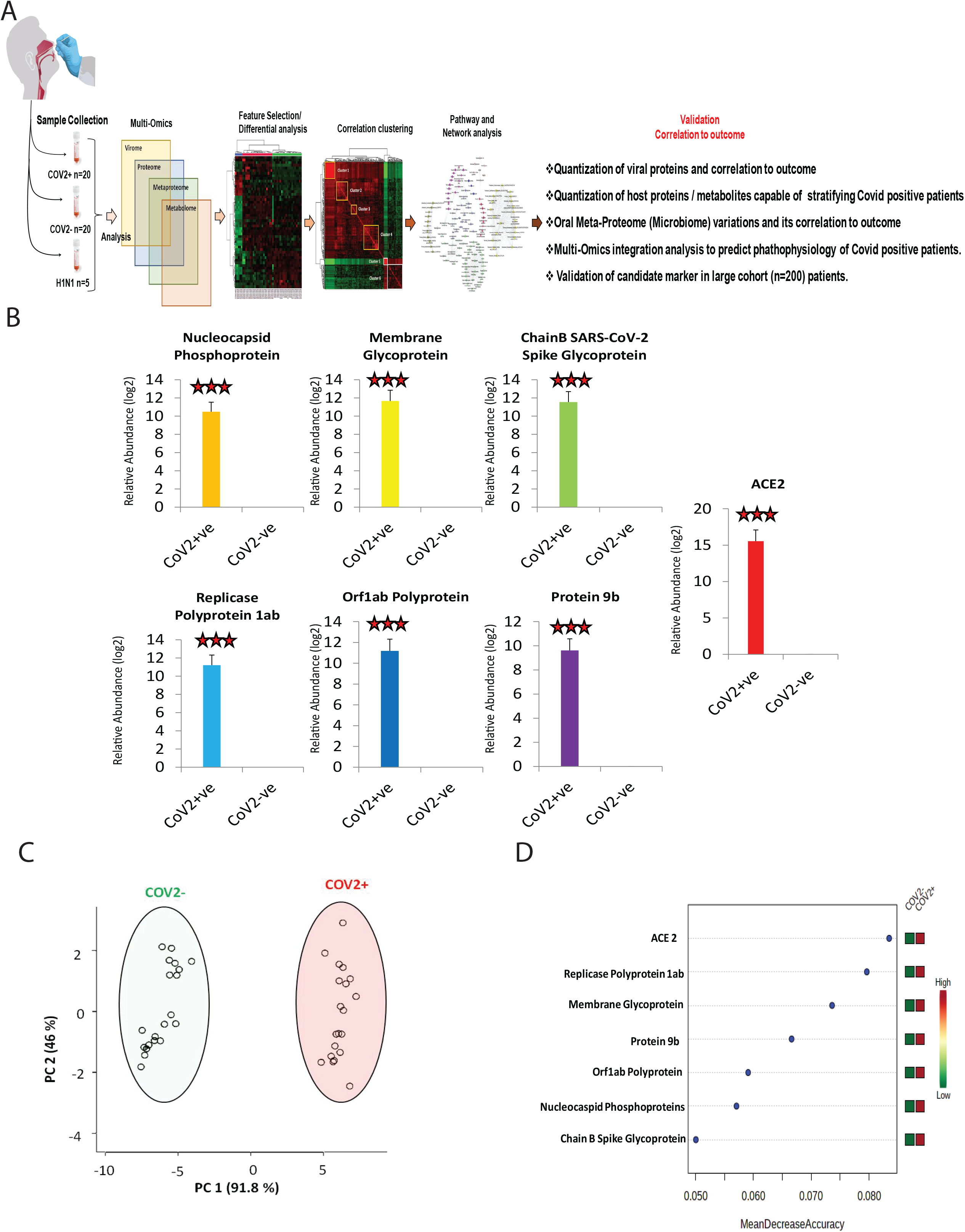
Quantitative virome profile of the respiratory specimen. **A:** Work flow for COVID-19 biomarker identification (discovery phase): Multi-omics (viromics, proteomics, metabolomics and metaproteomics) analysis was performed in 20 COVID-19 positive, 20 COVID-19 negative and 5 Influenza A H1N1 pdm 2009 positiverespiratory specimens. Differentially regulated viral/ host proteins, metabolites and bacterial peptides were identified which was subjected to random forest and AUROC analysis for identification of candidate for validation. Correlation clustering of the differentially expressed proteins, metabolites and peptides resulted in the identification of correlation clusters. Pathway analysis of the correlation clusters provided insight on the pathophysiology of SARS-COV2 infection. Finally the identified candidate indicator of COVID-19 was validated in 200 COVID suspect. This study attempted to answer the stated question. **B:** Log normalized abundance of 6 viral and 1 viral associated proteins identified in the respiratory specimen of COVID-19 positive as compared to COVID-19 negative patients. *** signifies p<0.05. **C:** Principle component analysis score plot showing segregation of COVID-19 positive patients from COVID-19 negative based on the identified Viral and associated proteins. **D:** Random forest analysis and mean decrease in accuracy plot showing the mean decrease in accuracy of the viral and associated proteins along with their expression status Red= upregulated and Green= downregulated in COVID-19 positive as compared to COVID-19 negative patients.

#### Diagnostic accuracy

Diagnostic accuracy was calculated using random forest analysis and the area under the receiver operating curve (AUROC) analysis showed that the mean decrease in the accuracy was highest for ACE2 followed by viral proteins as showed in Figure 1D, Supplementary Figure 1. Together, these analyses showed that there is significant increase in viral or viral linked protein expression in the respiratory specimens of COVID-19 positive patients compared to the negative patients and quantitation of viral/ virus associated proteins could be used for early diagnosis of SARS-CoV-2 infection.

### Proteome profile of the respiratory specimen could stratify COVID-19 positive patients

#### Identification of diagnostic proteins

To identify proteins having diagnostic /prognostic value and to have an insight in the pathogenesis of SARS-CoV-2 infection. Label free quantitative proteomics was performed (Supplementary Figure 2) on various study groups. Partial least square discriminant analysis (Figure 2A, Supplementary Figure 3) and unsupervised hierarchical clustering analysis (Figure 2B) clearly segregated COVID-19 positive from negative patients or H1N1 patients. Of the 1256 proteins identified, 132 were differentially expressed (DEP; 116 up- and 16 downregulated) in COVID-19 positive as compared to negative patients (FC>2, P<0.05, Supplementary Table 1, Supplementary Figure 4). Additionally, 166 DEP’s (117 up- and 49 downregulated) were identified in COVID-19 positive patients compared to H1N1 patients (FC>2, P<0.05, Supplementary Table 1, Supplementary Figure 4).

**Figure 2:**
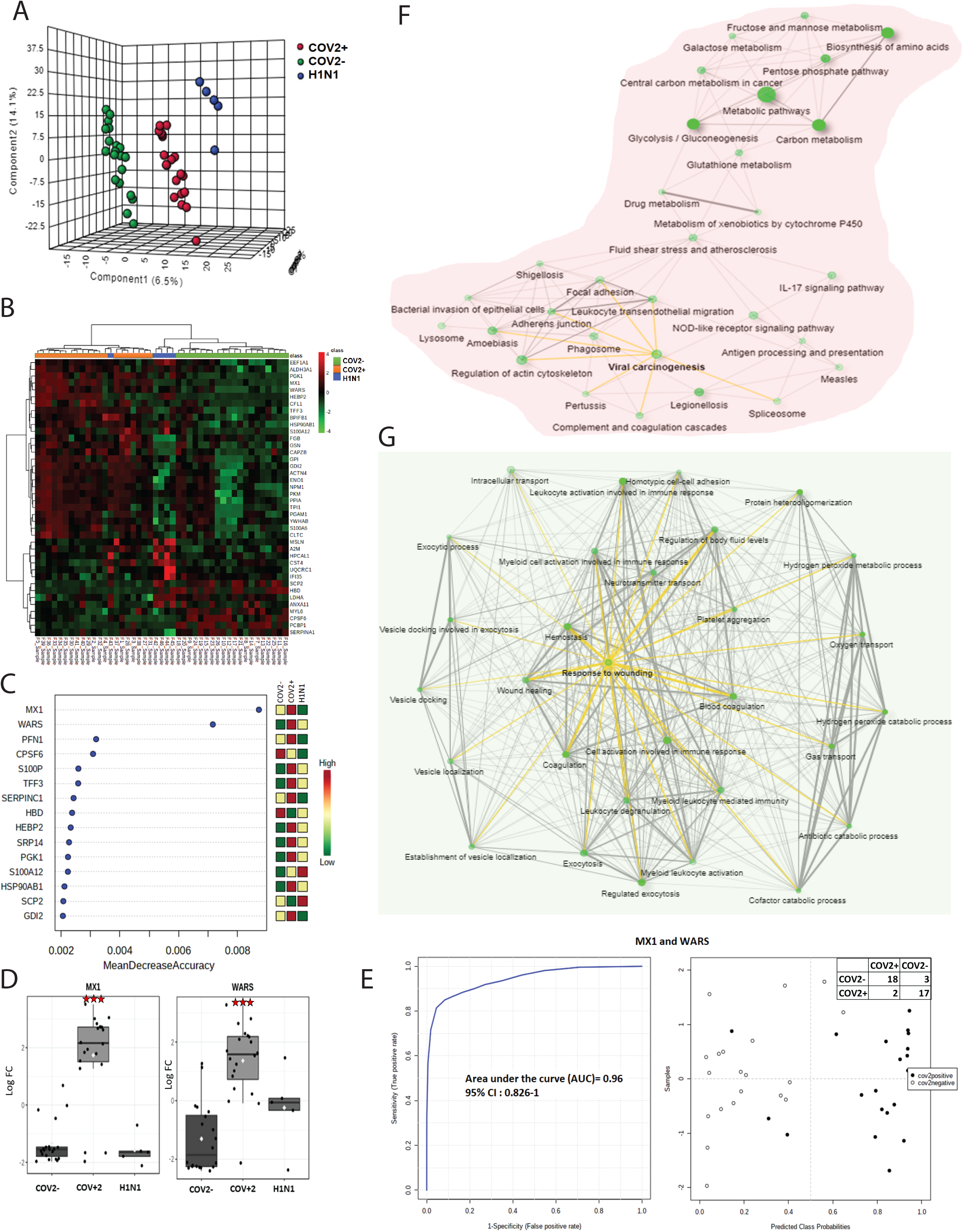
Respiratory specimen proteome profile insight on the pathophysiology of SARS-COV2 infection and stratifies COVID-19 positive patients. **A:** Partial least square discriminant analysis showing clear segregation of COVID-19 positive patients (Red dots) from COVID-19 negative (Green dots) and Influenza A H1N1 pdm 2009 positive patients (Blue dots) based on proteomic evaluation. **B:** Heat map and hierarchical cluster analysis of top 40 proteins (p<0.05) capable to segregate COVID-19 positive (orange bar) patients from COVID-19 negative (green bar) or Influenza A H1N1 pdm 2009 positive patients (blue bar). The expression is given as Red= upregulated, Green= downregulated and black= unregulated. **C:** Random forest analysis and mean decrease in accuracy plot showing the mean decrease in accuracy of the proteins along with their expression status Red= upregulated and Green= downregulated and yellow= unchanged in COVID-19 positive as compared to COVID-19 negative or Influenza A H1N1 pdm 2009 positive patients. **D:** Relative abundance (Log normalized) for MX1 and WARS showing significant increase in MX1 and WARS level in COVID-19 respiratory specimen as compared to COVID-19 negative or Influenza A H1N1 pdm 2009 positive cases (p-value**= p<0.05, ***<0.01). **E:** Joint AUROC analysis of MX1 and WARS documenting an AUC= 0.96 CI (0.82-1) p<0.05 along with prediction class probability score plot showing segregation of COV2 positive and COV2 negative. **F:** KEGG pathway analysis of 184 proteins up regulated in COVID-19 positive respiratory specimen as compared to COVID-19 negative or Influenza A H1N1 pdm 2009 positive Cases (FC>2; p<0.05, FDR>0.05). Darker nodes are more significantly enriched gene sets. Bigger nodes represent larger gene sets. Thicker edges represent more overlapped genes. Yellow edge show viral linked network **G:** KEGG pathway analysis of 60 proteins downregulated in COVID-19 positive respiratory specimen as compared to COVID-19 negative or Influenza A H1N1 pdm 2009 positive cases (FC>2; p<0.05, FDR>0.05). Darker nodes are more significantly enriched gene sets. Bigger nodes represent larger gene sets. Thicker edges represent more overlapped genes. Three clusters of edges can be identified: (a) gas transport and oxygen transport cluster (b) wound healing and inflammation cluster and (c) vesicle transport cluster. Yellow edge show wound healing linked network.

**Table 1:**
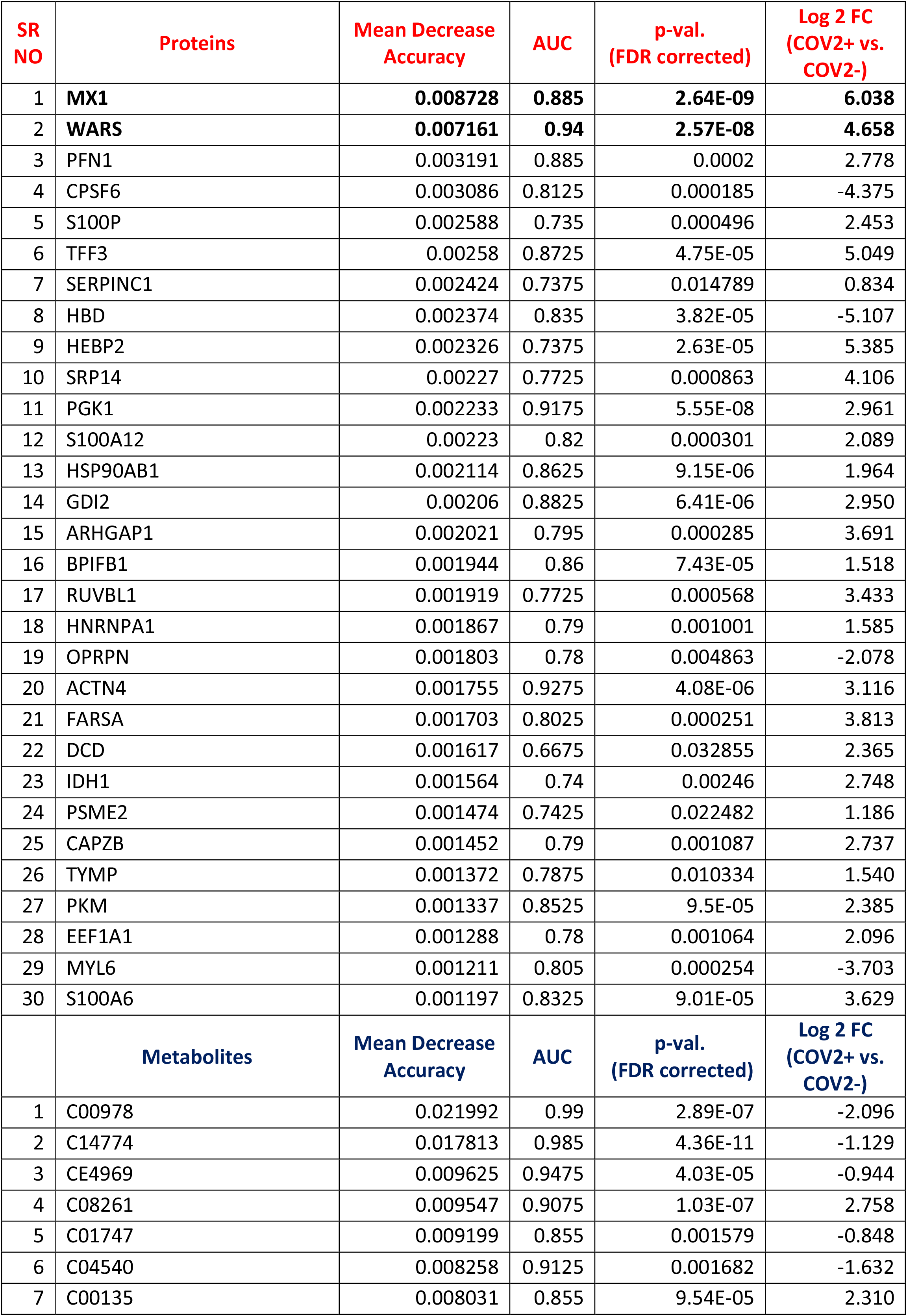
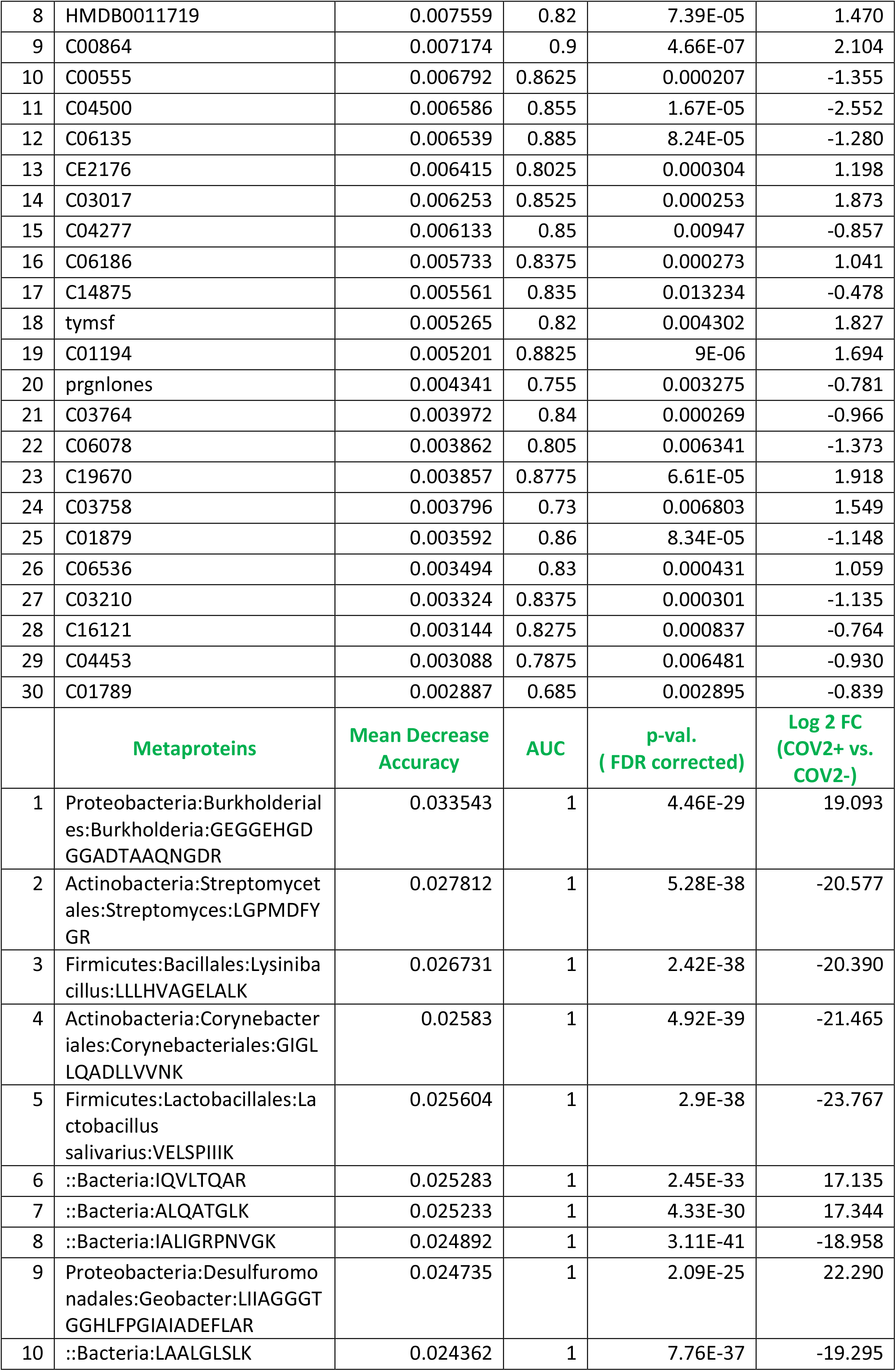
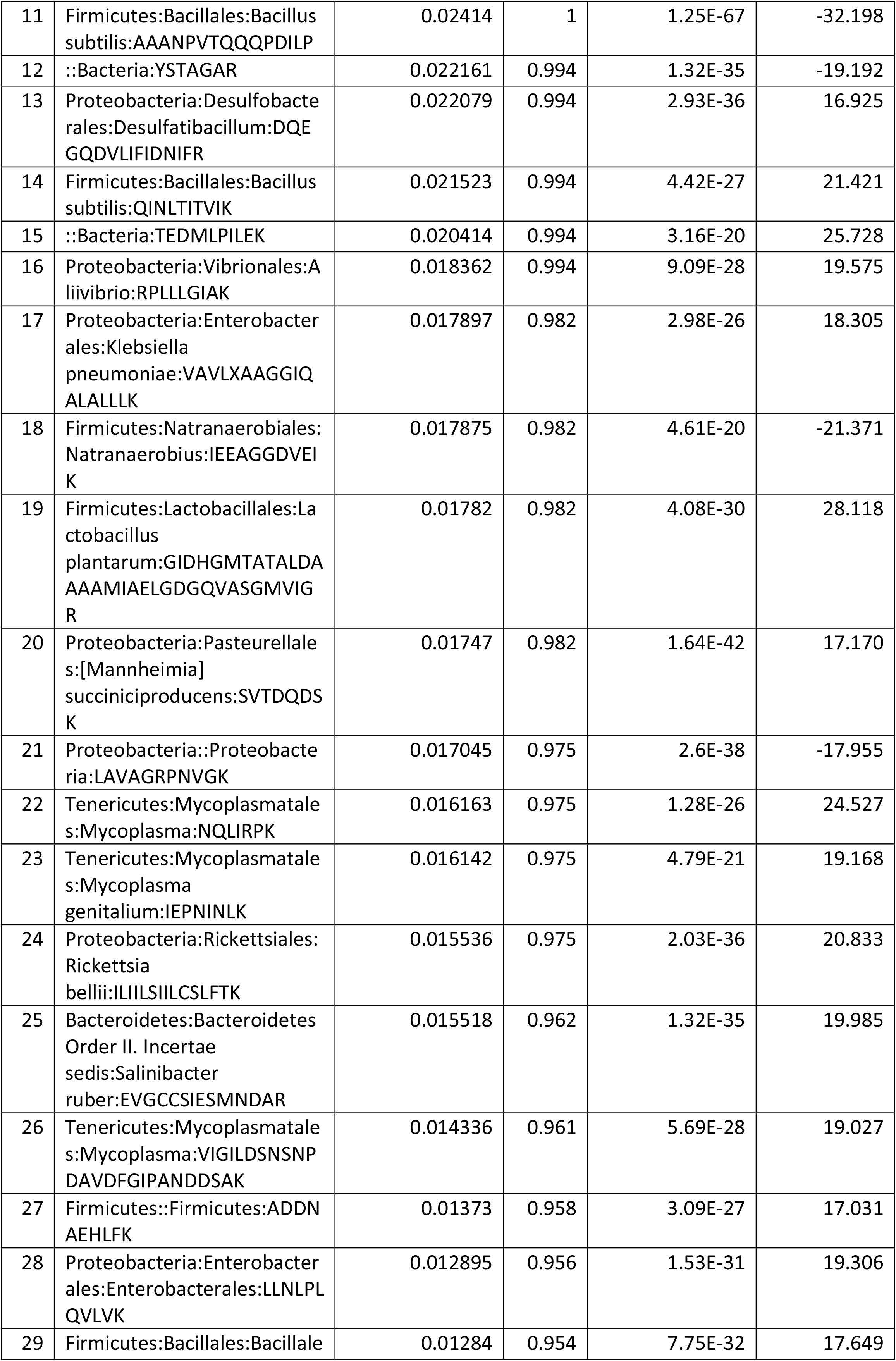
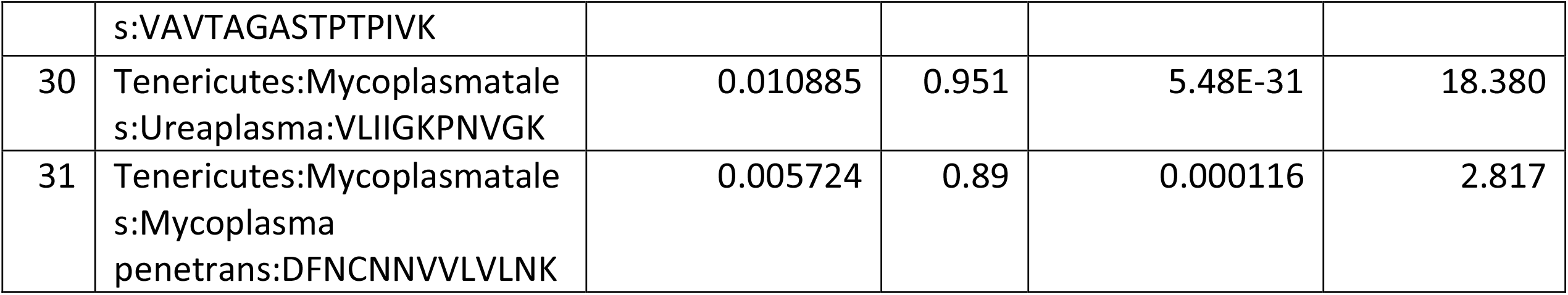
Biomarker panel identified from Multi-omics analysis

#### Diagnostic accuracy

Amongst the identified DEP’s, mean decrease in the accuracy (calculated by random forest; 1000 trees) was highest for MX1 (MX Dynamin like GTPase 1) and WARS (Tryptophan--tRNA ligase) making them the most important proteins for segregating COVID-19 positive patients from negative or H1N1 patients (Figure 2C, 2D). The diagnostic efficiency (AUROC) of MX1 was 0.895 (0.746-1) and WARS was 0.948 (0.85-1) for COVID-19 positive detection (p<0.05; Supplementary Figure 8). Combined diagnostic efficiency of MX1 and WARS was found to be 0.96 (0.826-1) and showed clear segregation of COVID-19 positive from COVID-19 negative patients (Figure 2E).

#### Biological relevance

Proteins significantly upregulated in COVID-19 positive patients were enriched for pathways linked to viral carcinogenesis, interferon signaling, IL-2 signaling, IL-3, IL-5 and GMCSF signaling linked to monocyte activation and inflammation, complement activation, immune activation (IL-17 signaling, NOD-like receptor signaling, neutrophil and leukocyte activation, transendothelial migration and antigen presentation), bacterial invasion of epithelial cells, fluid shear stress (exocytosis, vascular transport and cell secretions), Drug, xenobiotic, glutathione and glucose metabolism (Figure 2F, KEGG pathways; FDR<0.05, Supplementary Figure 5, Supplementary Figure 6). Proteins significantly downregulated in COVID-19 positive patients were enriched for pathways linked to wound healing, haemostasis, regulation of body fluids, exocytosis, platelet aggregation, gas transport, oxygen transport (heamoglobin binding, haptoglobin binding), hydrogen peroxide and cofactor catabolic/metabolic process, and vesicular transport (Figure 2G, KEGG pathways; FDR<0.05, Supplementary Figure 7).

Together these findings showed that COVID-19 positive patients have virus mediated hyper immune activation involving monocytes and neutrophils, deregulated oxygen transport, increased fluid shear stress, bacterial invasion of the epithelial cells and glucose metabolism. Further, proteins such as MX1 (MX Dynamin Like GTPase 1) and WARS (Tryptophan--tRNA ligase) are regulated by SARS-COV2 and could be validated for a probable indicator of COVID-19 diagnosis.

### Metabolic phenotype of the respiratory specimen is predictive of SARS-CoV-2 infection

#### Identification of diagnostic metabolites

SARS-CoV-2 infection could change the metabolic phenotype of respiratory specimen; recently a distinctive plasma metabolomic profile associated with SARS-CoV-2 infection and severity is also reported (Shen et al., 2020). Thus, viral pathogenesis (hyper immune activation) seen in SARS-CoV-2 infection could be attributed to a change in the respiratory metabotype. We evaluated and compared the respiratory metabolome in COVID-19 positive, negative and H1N1 patients (Supplementary Figure 9). Metabolomic analysis of the respiratory specimen identified 106 differentially expressed metabolites (DEMs) in COVID-19 positive patients (53up- and 53 downregulated) as compared to negative (Figure 3A, FC>1.5, P<0.05, Supplementary Figure 10, Supplementary Table 2). In addition, 274 DEM’s were identified when COVID-19 positive patients were compared to H1N1 patients (Figure 3A, FC>1.5, P<0.05, Supplementary Figure 10, Supplementary Table 2). Partial least square discriminant analysis (PLS-DA) followed by unsupervised clustering analysis clearly segregated COVID-19 positive from COVID-19 negative or H1N1 cases (Figure 3B, 3C, Supplementary Figure 11).

**Figure 3:**
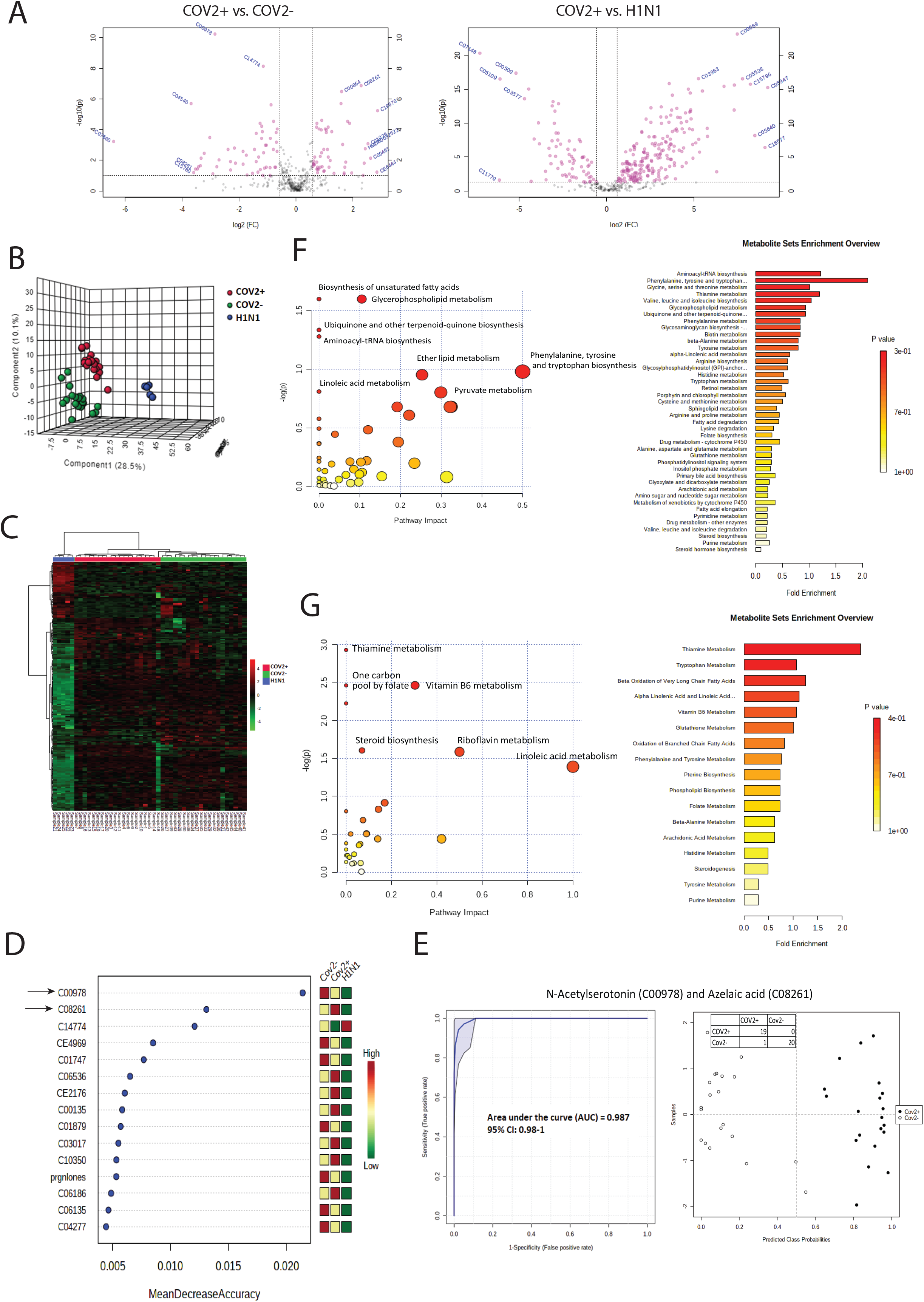
Metabolic phenotype of the respiratory specimen is predictive of SARS-COV2 infection. **A:** Volcano plot showing differentially expressed metabolites in COVID-19 positive vs. COVID-19 negative respiratory specimen and COVID-19 positive vs. Influenza A H1N1 pdm 2009 positive specimen. Pink dots are significant (p<0.05, FC>1.5) **B:** Partial least square discriminant analysis showing clear segregation of COVID-19 positive patients (Red dots) from COVID-19 negative (Green dots) and Influenza A H1N1 pdm 2009 positive cases patients (Blue dots) based on metabolomic estimations. **C:** Heat map and hierarchical cluster analysis of the metabolites identified in the study (p<0.05) show clear segregation of COVID-19 positive patients (Red bar) from COVID-19 negative (Green bar) or Influenza A H1N1 pdm 2009 positive cases patients (Blue bar). The expression is given as Red= upregulated, Green= downregulated and black= unregulated. **D:** Random forest analysis and mean decrease in accuracy plot showing the mean decrease in accuracy of the Metabolites along with their expression status Red= upregulated and Green= downregulated and yellow= unchanged in COVID-19 positive as compared to COVID-19 negative or Influenza A H1N1 pdm 2009 positive patients. **E:** Joint AUROC analysis of N-acetylserotonin (C00978) and azelaic acid (C08261) and documenting an AUC= 0.987 CI (0.98-1) p<0.05 along with prediction class probability score plot showing segregation of COV2 positive and COV2 negative. **F:** Pathway analysis and metabolite set enrichment analysis (KEGG) for the upregulated metabolites (FC> 1.5, p<0.05) in COVID-19 positive respiratory specimen as compared to COVID-19 negative or Influenza A H1N1 pdm 2009 positive specimen. **G:** Pathway analysis and metabolite set enrichment analysis (KEGG) for the downregulated metabolites (FC> 1.5, p<0.05) in COVID-19 positive respiratory specimen as compared to COVID-19 negative or Influenza A H1N1 pdm 2009 positive specimen.

#### Diagnostic accuracy

Amongst the DEM’s, mean decrease in the accuracy (calculated by random forest; 1000 trees) was found to be highest for N-acetylserotonin (C00978) and azelaic acid (C08261) making them the most important metabolites capable of segregating COVID-19 positive patients (Figure 3D). The diagnosis efficiency (AUROC) of N-acetylserotonin was 0.998 (0.972-1) and azelaic acid were 0.941 (0.85-1) for COVID-19 positive detection (p<0.05; Supplementary Figure 12), with combined diagnostic efficiency of 0.987 (0.98-1) to segregate SARS-CoV-2 positive from negative patients (Figure 3E).

#### Biological relevance

Metabolites significantly upregulated in COVID-19 positive patients were enriched for pathways linked to biosynthesis of unsaturated fatty acids, glycerophospholipid metabolism, ubiquinone/terpenoid-quinone biosynthesis, aminoacyl-tRNA biosynthesis, amino acid metabolism including phenylalanine, tyrosine and tryptophan biosynthesis (Figure 3F). Metabolites significantly downregulated in COVID-19 positive patients were enriched for pathways linked to thiamine metabolism, one carbon pool by folate, vitamin B6 metabolism, riboflavin metabolism and steroid biosynthesis (Figure 3G).

Together these results suggest that respiratory specimen of COVID-19 positive patients are rich in oxidative and inflammatory metabolite including tryptophan and derivative (3-hydroxyanthranilate, cinnavalininate, N-methyltryptamine), histidine, fatty acids, glycerolipids, aminoacyltRNA and have significantly lower levels of anti-inflammatory steroids or vitamins. In addition, metabolites such as N-acetylserotonin (C00978) and azelaic acid (C08261) could segregate COVID-19 positive patients and warrant validation in larger cohort.

### SARS-COV2 infection modulates Metaproteome (Microbiome) profile of respiratory specimen with possibility to predict COVID-19 positivity

#### Identification of diagnostic metaproteome

Often SARS-CoV-2 infection precedes bacterial co-infection and is linked with longer duration and more severe infection (Bengoechea and Bamford, 2020; Gu and Korteweg, 2007). SARS-CoV-2 is also known to modulate the gut microbiota and is associated to immune cell activation and severe outcome (Dhar and Mohanty, 2020). We hypothesized that change in naso-pharyngeal/ oropharyngeal microbiome (likely source of lung microbiome) due to SARS-CoV-2 infection could be reflective of altered pathogenesis and help in diagnosis of COVID-19 diseases. To explore this, metaproteome analysis was performed in the respiratory specimen of the study group (Supplementary Figure 13). Principle component analysis along with unsupervised clustering analysis showed clear segregation of the COVID-19 positive from other groups (Figure 4A, 4B). At the phylum level Proteobacteria and Tenericutes were increased, corroborating to an increase in alpha diversity seen in COVID-19 positive respiratory specimen (Figure 4C, 4D, p<0.05). Linear discriminating analysis showed that COVID-19 respiratory specimen has significantly increased bacterial peptide linked to the (phylum: LCA); Bacteroidetes: Bacteroidetes Order II. Incertae sedis, Firmicutes: Bacillales, Bacillus subtilis, Lactobacillus plantarum, Proteobacteria: Burkholderiales, Desulfobacterales, Desulfuromonadales, Enterobacterales, Klebsiella pneumonia and others, Tenericutes: Mycoplasma genitalium, Mycoplasma mobile and others whereas bacterial peptide linked to Actinobacteria: Corynebacteriales, Streptomycetales, Firmicutes: Bacillus subtilis, Lysinibacillus, Lactobacillus salivarius and others, Proteobacteria : Gamma proteobacteria, pseudomonadales and others and Tenericutes:Mycoplasmatales were significantly reduced (p<0.05; Figure 4E, Supplementary Table 3) as compared to other groups.

**Figure 4:**
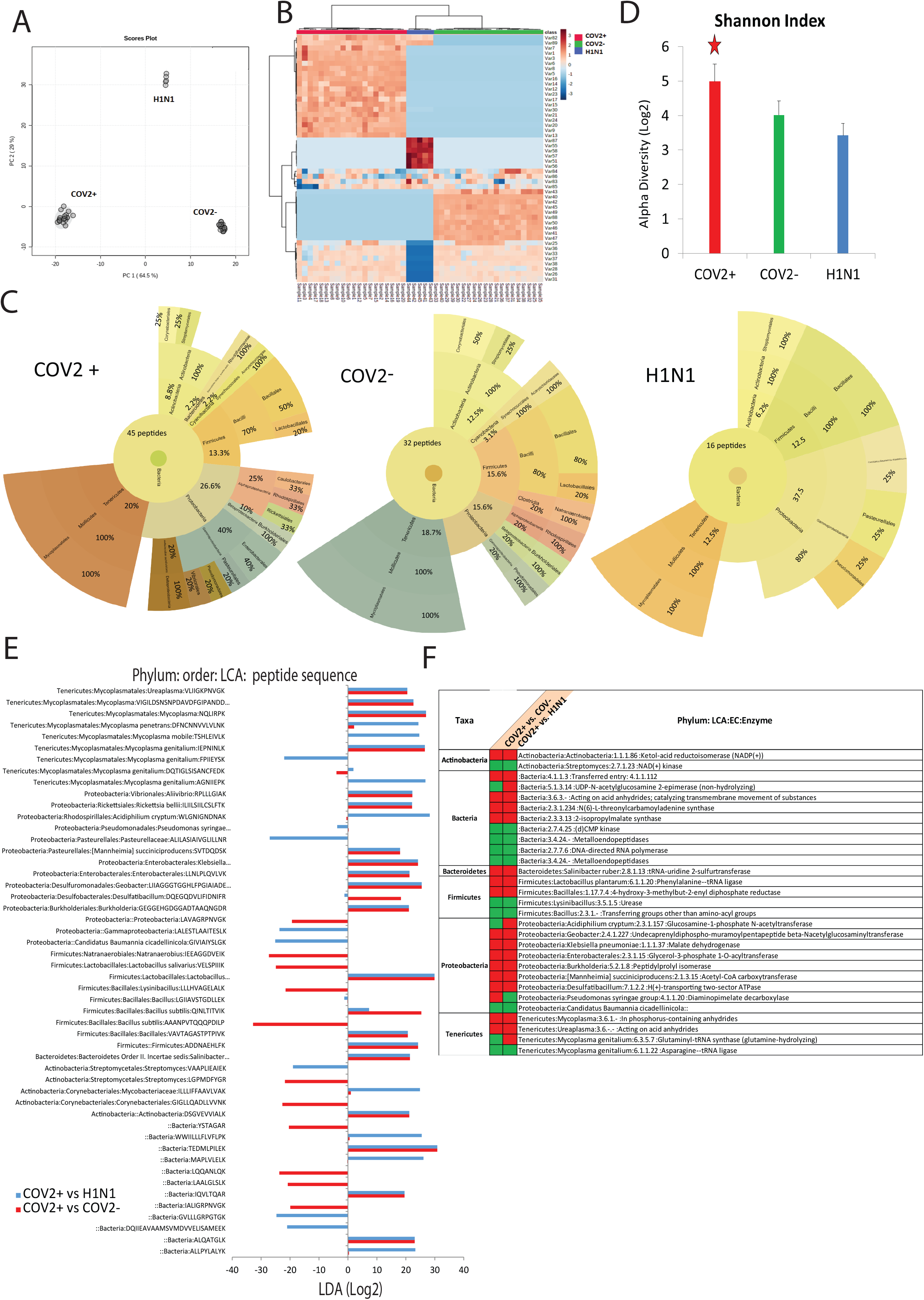
Respiratory specimen Metaproteome (Microbiome) could segregate COVID-19 positive patients. **A:** Principle component analysis (PCA) showing clear segregation of COVID-19 positive patients from COVID-19 negative and Influenza A H1N1 pdm 2009 positive patients based on metaproteins estimations. **B:** Heat map and hierarchical cluster analysis of the metaproteins identified in the study (p<0.05) show clear segregation of COVID-19 positive patients (Red bar) from COVID-19 negative (Green bar) or Influenza A H1N1 pdm 2009 positive patients (Blue bar). The expression is given as Dark brown= upregulated, Blue= downregulated and white= unregulated. **C:** Sunburst plot representative of microbial population difference (phylum: order: family) in COVID-19 positive respiratory specimen as compared to COVID-19 negative or Influenza A H1N1 pdm 2009 positive cases. Total no of peptides and their percent distribution identified in provided in each sunburst plot **D:** Alpha diversity index calculated based on the relative abundance of bacterial peptides identified in COVID-19 positive respiratory specimen as compared to COVID-19 negative or Influenza A H1N1 pdm 2009 positive cases (p-value *=<0.05). **E:** Linear discriminant analysis showing log2 levels of the identified bacterial peptides in the respiratory specimen of COVID-19 positive compared to COVID-19 negative (Red) and COVID-19 positive compared to Influenza A H1N1 pdm 2009 positive cases (Blue). **F:** Enzyme coded by the bacterial peptides and their abundance in COVID-19 positive compared to COVID-19 negative and COVID-19 positive compared to Influenza A H1N1 pdm 2009 positive cases Expression of the enzyme is provided Red shows upregulated and Green shows down regulated.

#### Diagnostic accuracy

Amongst the differentially expressed meta-proteins (DEMPs), mean decrease in the accuracy (random forest) was highest for Bacillus subtilis (Variable 82), and Burkholderiales (Variable 30) making them the most important bacterial LCA which could segregate COVID-19 positive from negative or H1N1 cases (AUROC>0.99; p<0.05, Supplementary Figure 14 and 15).

#### Biological relevance

Functionality of the COVID-19 positive respiratory microbiome highlighted significant increase in enzymes linked to pantothenate and CoA biosynthesis in actinobacteria (EC1.1.1.86), trna-uridine 2-thiolation in bacteroidetes (EC2.8.1.13:), aminoacyl-trna biosynthesis and terpenoid backbone biosynthesis in fermicutes (EC6.1.1.20, EC1.17.7.4), amino-nucleotide sugar metabolism, peptidoglycan biosynthesis, fatty acid biosynthesis and glycerophospholipid metabolism, energy metabolism and oxidative phosphorylation (EC2.3.1.157, EC2.4.1.227, EC1.1.1.37, EC2.3.1.15, EC2.1.3.15, EC7.1.2.2, EC4.1.1.20) in proteobacteria and aminoacyl-trna biosynthesis in tenericute (EC3.6.1, EC6.3.5.7) a clear indication of bacterial pathogenic thrust in presence of CoV2. Whereas enzyme linked to nicotinate and nicotinamide metabolism (EC2.7.1.23) in actinobacteria, arginine biosynthesis (EC3.5.1.5) in fermicutes, and Aminoacyl-tRNA biosynthesis linked to aspartate and asparagine metabolism in tenericute were decreased (Figure 4F).

Together these results suggest that SARS-CoV-2 infection modulates the oropharyngeal microbiome and associated function which correlates with COVID-19 pathophysiology and could help in COVID-19 diagnosis.

### Multi-omics integration analysis delineates pathways linked to SARS-CoV-2 pathogenesis

We hypothesized that change in the viral proteome (viral replication) should reflect a corresponding/analogues change in the host proteome, metabolome and metaproteome (microbiome). For example, an increase in viral proteome or bacterial proteome should recapitulate a corresponding increase in proteins or metabolites associated pathways and members of known pathways should co-cluster. This multi-omics integration analysis could help in classification of class membership of metabolites, proteins, familial enzyme pathways or novel enzyme reaction models associated to change in viral proteome and pathogenesis. To explore this, global cross-correlation and hierarchal clustering analysis were performed for differentially expressed viral proteome (DEVP: 7), differentially expressed proteins (DEP: 132), differentially expressed metaproteome (DEMP: 33) and differentially expressed metabolites (DEM: 106) in COVID-19 positive patient as compared to COVID-19 negative (Figure-5A). Principle component analysis recapitulates the distinction between COVID-19 positive and negative patients (Figure 5B). Correlation analysis followed by hierarchical clustering (r^2^>0.5, p<0.05) showed that viral proteins significantly correlates with bacterial and host proteins or metabolites and identified 6 clusters (Figure-5C). Mean intensity of cluster 1 and cluster 5 was significantly different in COVID-19 positive patients as compared to negative (p<0.05, Figure 5D). Pathway analysis for the proteins /metabolites linked with each cluster (Figure 5E) showed that increase in the viral proteome: cluster 1 was associated with significant increase in metaproteome (Bacterial taxa), proteins/metabolites linked to viral life cycle, regulation of angiogenesis, vasculature development, protein processing and maturation, beta-alanine metabolism, pantothenate and CoA biosynthesis and glycosylphosphatidylinositol (GPI) anchor biosynthesis (r^2^>0.5, p<0.05, Figure 5E, supplementary table 4). In addition decrease in the bacterial taxa: cluster 5 was linked with significant decrease in oxidative phosphorylation, thermogenesis, beta oxidation of fatty acids, and arachidonic acid metabolism (r^2^>0.5, p<0.05, Figure 5E, supplementary table 4). This analysis identifies the underlying metabolic variations associated with naso-pharyngeal/ oropharyngeal dysbiosis under CoV-2 infection. Over all, these results establish a linear and direct relationship between viral proteome, bacterial metaproteome, host proteome, and metabolome in COVID-19 positive respiratory specimen.

**Figure 5:**
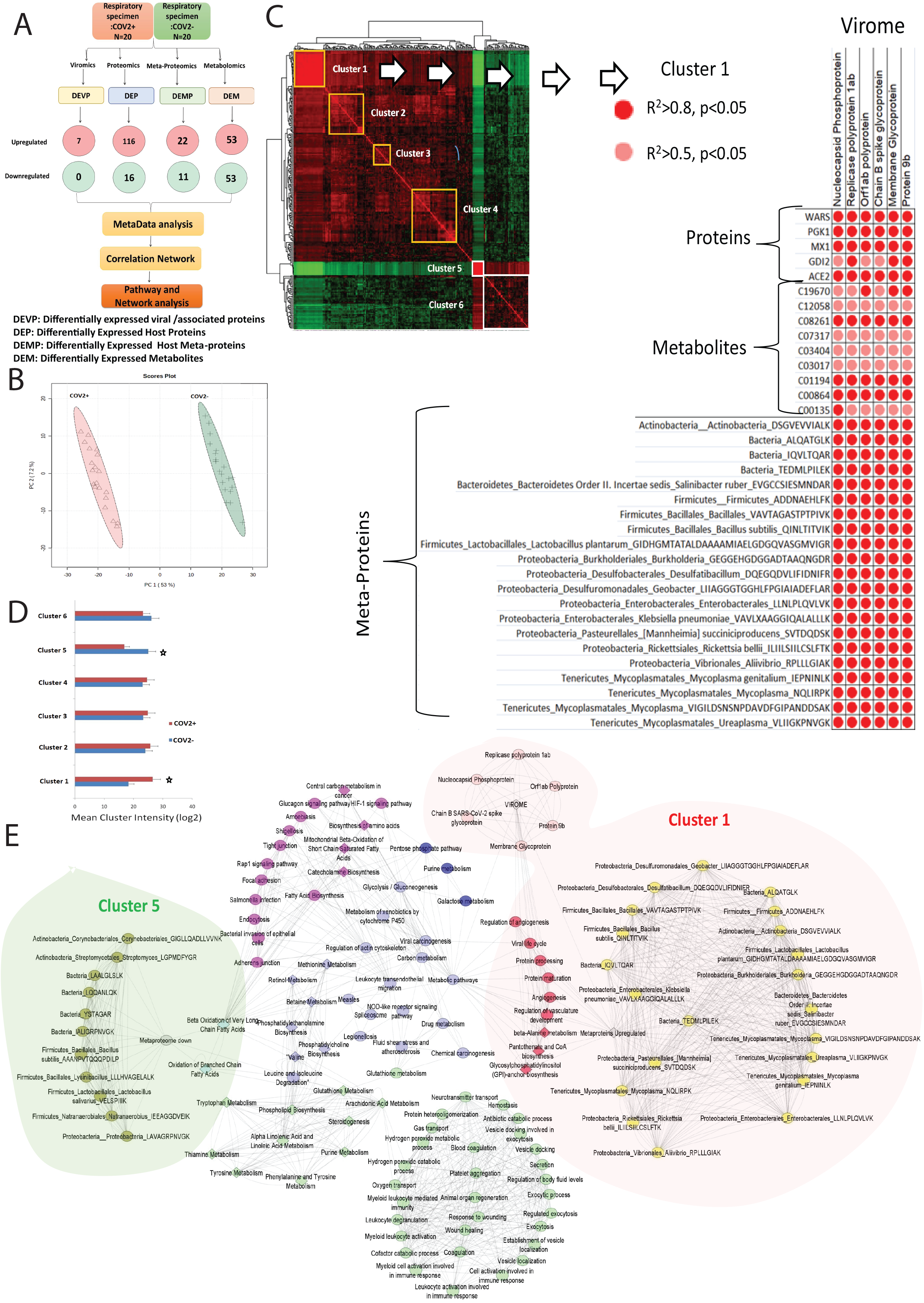
Respiratory multi-omics cross correlation delineate pathways linked to SARS-COV2 persistence and pathogenesis. **A:** Schematic representation of the integration analysis performed in this study. DEVP: differently expressed viral /associated proteins, DEP: differently expressed host proteins, DEMP: differently expressed host meta-proteins, DEM: differently expressed metabolites were cross correlated to identify viral regulatory network. **B:** Principle component analysis (PCA) showing clear segregation of COVID-19 positive patients from COVID-19 negative based on the multi-omics profile. **C:** Global cross correlation analysis led to identification of 6 clusters (4 upregulated marked in yellow and 2 down regulated marked in white). Details of cluster 1 is shown adjacent which show viral protein along with bacterial peptides, proteins and metabolites correlation with each other. Each correlation is represented with a dot where red dot=r2>0.8, p<0.05 and pink dot=r2>0.5, p<0.05 respectively. **D:** Mean cluster intensity is shown Cluster 1 and Cluster 5 are significantly different in COVID-19 positive compared to COVID-19 negative patients (p<0.05). **E:** Global correlation map is shown for the six cluster identified; Cluster1: RED, Cluster2: pink, Cluster3: blue, Cluster4: purple, Cluster5: light blue, Cluster6: light green. Viral proteins are shown in light pink and bacterial peptide is shown in yellow (upregulated) and olive green (downregulated). The most important cluster1 and cluster5 are highlighted. Connecting line are representing correlation r2>0.5 p<0.05 respectively.

### Respiratory multi-omics correlates with viraemia, show increase in monocyte, platelet and immune activation and segregate non symptomatic COVID-19 patients

High viral load inversely relates to the CT values (RT-PCR analysis) (Lirong Zou nejm 2020). We wanted to identify which parameters of the respiratory multi-omics data correlates with the viraemia of COVID-19 positive patients. For this the identified DEVP, DEP, DEMP and DEM values were correlated with the CT values of the COVID-19 positive patients. A total of 54 proteins and/or metabolites showed significant correlation with the CT values of COVID-19 positive patients (Figure 6A, Supplementary Table 5). Proteins and metabolites linked to energy metabolism (Pentose phosphate pathway, Glycolysis), Glycerophospholipid metabolism, Purine metabolism and inflammation (IL-17 signaling pathway) correlated inversely with the CT values and were increased with increase in Viraemia (R^2^>0.5, p<0.05, Figure 6A, Supplementary Table 5).

**Figure 6:**
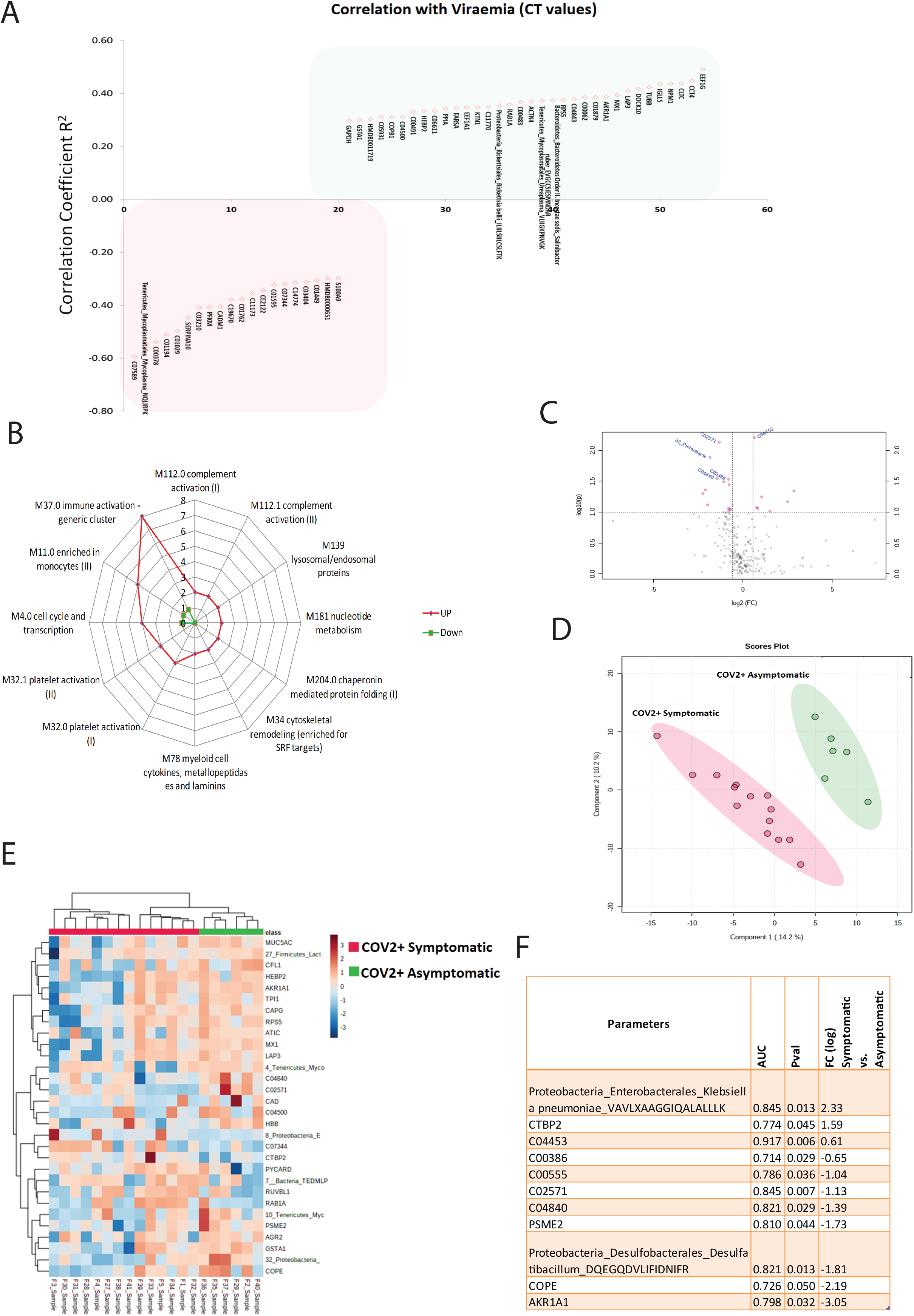
Respiratory multi-omics correlates with viraemia, show increase in monocyte, platelet and immune activation and segregate non symptomatic COVID-19 patients. **A:** Correlation plot showing top 54 proteins and metabolites which showed significant correlation with the CT values of COVID-19 positive patients. Green highlight on the molecules are positively correlated to the CT values and Red color highlight on the molecules denotes negative correlation with the CT values (RT-PCR). **B:** BTM enrichment analysis of respiratory proteome identifies immune clusters represented as web plot showing the number of proteins present in each module, red= upregulated in COVID-19 respiratory specimen and green= downregulated in COVID-19 respiratory specimen. **C:** Volcano plot showing differentially expressed multi-omics data in asymptomatic COVID-19 as compared to symptomatic COVID-19 respiratory specimen. Pink dots are significant (p<0.05, FC>1.5) **D:** Partial least square discriminant analysis showing clear segregation of symptomatic COVID-19 patients (Red dots) from asymptomatic COVID-19 patients (Green dots) based on Multi-omics estimations. **E:** Heat map and hierarchical cluster analysis of the multi-omics identified (p<0.05) show clear segregation of COVID-19 symptomatic (Red bar) from asymptomatic COVID-19 (Green bar) patients. The expression is given in the range as Dark brown= upregulated, Blue= downregulated and white= unregulated. **F:** Area under the receiver operating curve analysis along with fold change and significance of bio-molecules capable of segregating COVID-19 symptomatic from asymptomatic COVID-19 patients.

Next we analyzed the respiratory specimen for immune imprints by enriching the differentially expressed proteome (DEP) on the blood transcription module (BTM) space (Li et al., 2017). The upregulated proteins were enriched for modules associated to immune activation generic cluster, enriched in monocytes, cell cycle and transcription and platelet activation module I and II (> 3 genes, Figure 6B, Supplementary table 6) and showed direct correlation with the metabolites linked to arginine biosynthesis, cysteine and methionine metabolism and Aminoacyl-tRNA biosynthesis. Correlation of immune, monocyte and platelet clusters with metabolic pathways highlights the inflammatory and activated state of these cells in COVID-19 respiratory specimen

Next we explored the efficacy of the multi-omic profile in segregating asymptomatic COVID-19 positive patient. Of the 271(DEP, DEMP and DEM) a total of 20 variables could significantly differentiate COVID-19 asymptomatic patients (Figure 6C). Partial least square discriminate analysis (PLS-DA) along with unsupervised clustering analysis showed clear distinction between symptomatic and asymptomatic COVID-19 patients (Figure 6D). The diagnostic efficiency of AKR1A1 (Aldo-keto reductase family 1 member A), COPE (Coatomer subunit epsilon), PSME2 (Proteasome activator complex subunit 2) and C02571 (L-Acetyl Carnitine) was highest for segregation of asymptomatic COVID-19 patients while symptomatic COVID-19 patients showed significant increase in Klebsiella pneumoniae, CTBP2 (C-terminal-binding protein 2) and 4-alpha-Methyl-5-alpha-cholest-7-en-3-one (Figure 6E). Together these results show that multi-omics profile of COVID-19 positive respiratory specimen correlates with viraemia, provide an insight on immune activation and could also aid in the identification of asymptomatic COVID-19 patients.

### Multi-omics analysis of the respiratory specimen validates MX1 and WARS as candidate indicator of COVID-19 diagnosis

#### Validation

Multi-omics analysis led to the identification of a panel of molecules (Table 1) which could significantly segregate COVID-19 positive patients from negative or H1N1 cases. Of them we cherry picked MX1 (MX Dynamin like GTPase 1) and WARS (Tryptophan--tRNA ligase) as they were the most important proteins which could segregating COVID-19 positive patients. Expression of MX1 and WARS was evaluated in a larger cohort of 200 COVID-19 suspects. COVID-19 positive patients showed significant increase in MX1 and WARS expression as compared to COVID-19 negative (p<0.05, Figure 7A). The diagnostic efficiency of MX1 was 92% (AUROC=0.92(0.863-0.957)) and WARS was 86% (AUROC=0.867(0.781-0.944)) for segregation of COVID-19 positive patients (p<0.05, Figure 7B). MX1 independently showed a predictive accuracy of 83% (1000 permutation) in diagnosis of COVID-19 positive patients (Figure 7C, Supplementary Figure 16). MX1 >30pg/ml sensitivity (82.5% CI(73.7%-89.3%))- specificity(82.6% (73.76%-89.3%)) in combination with WARS >25ng/ml sensitivity (72.3% CI(65.7%-82.3%))- specificity(76.6% (66.6%-85.3%)) showed a diagnostic efficiency of 94% (AUROC=0.948 CI(0.911-0.977)) and a predictive accuracy of 86% (1000 permutation) in segregating COVID-19 positive from COVID-19 negative patients (Figure 7D, Figure 7E, Supplementary Figure 17). Together these results potentiate the utility of MX1 and WARS in segregation and diagnosis of COVID-19 patients.

**Figure 7:**
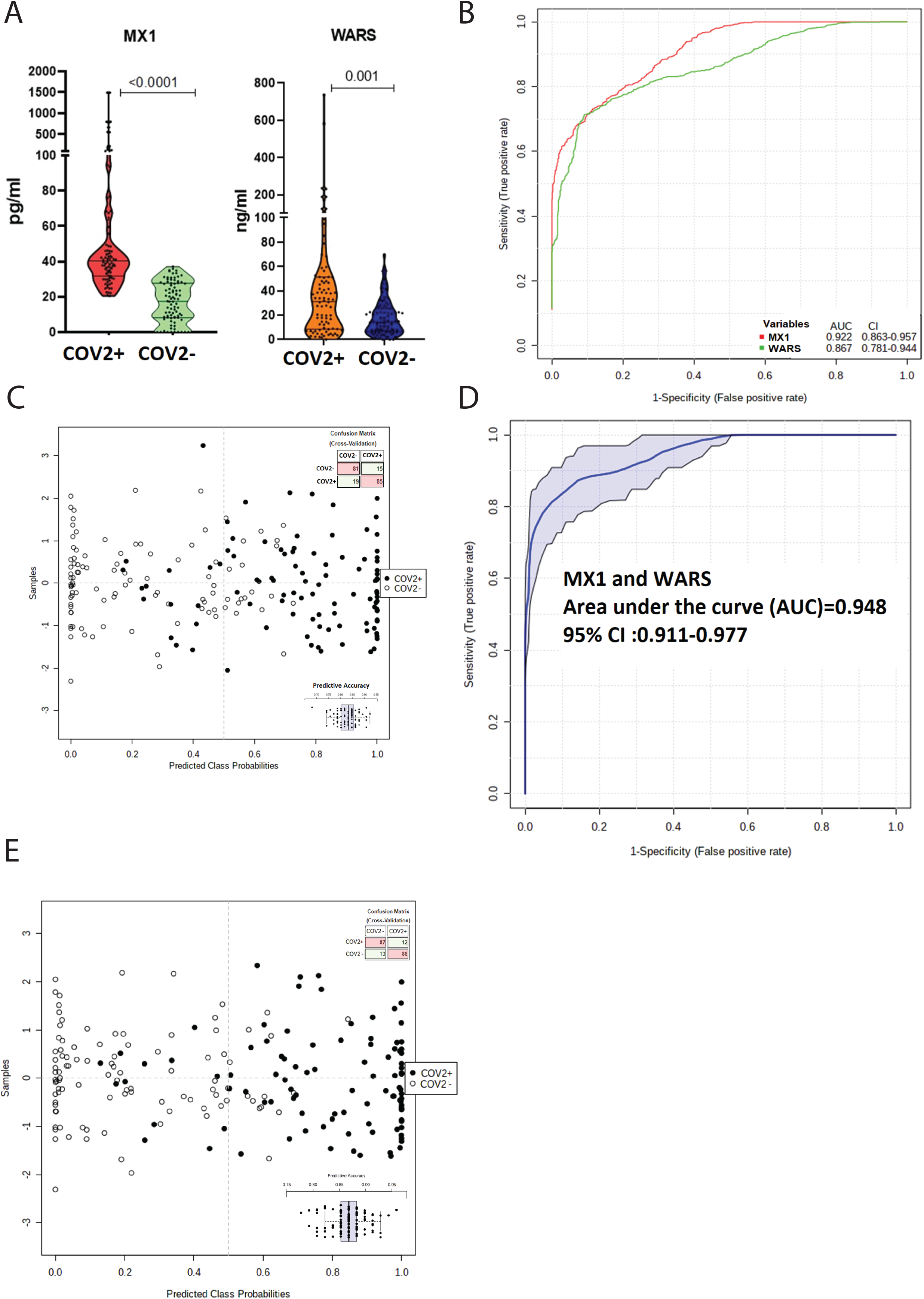
Estimation of MX1 and WARS as candidate indicators for COVID-19 diagnosis. **A:** Quantitative assessment of MX1 and WARS in the respiratory specimen of 200 COVID-19 specimen show significant increase in MX1 and WARS levels in COVID-19 positive as compared to COVID-19 negative (FC> 2, p<0.05) **B:** Multivariate area under the receiver operating curve analysis show significant AUC of MX1=0.922 CI (0.863-0.957) and WARS =0.867 CI (0.781-0.944). **C:** Prediction class probability for MX1 alone show clear segregation of COVID-19 positive from COVID-19 negative patients with a predictive accuracy of 84%. **D:** Area under the receiver operating curve analysis for MX1 and WARS together show AUC=0.948 CI (0.911-0.977) p<0.05. **E:** Prediction class probability for MX1 and WARS together show clear segregation of COVID-19 positive from COVID-19 negative patients with a predictive accuracy of 86%.

## Discussion

In the present study, the molecular profile of the respiratory specimen was analyzed for the characterization of baseline molecular determinants which could be used as putative candidate for SARS-CoV-2 diagnosis. This was complemented with global proteome (viral and host), metaproteome and metabolome analysis of the respiratory specimens followed by analysis of the regulatory network. Multi-omics integrated analysis helped us in the characterization of a panel of biomolecules capable of stratifying COVID-19 positive patients but also provided insight on the pathogenesis of SARS-CoV-2 infection. Of the panel, we validated the expression of MX1 (MX Dynamin like GTPase 1) and WARS (Tryptophan--tRNA ligase) in the respiratory specimen of 200 COVID-19 suspects and showed that increase in their expression corresponds to SARS-CoV-2 infection positivity. These molecules could be used as a putative candidate for SARS-CoV-2 infection monitoring or modulation/therapeutic targets.

SARS-CoV-2 is known to mediate infection by entry into the cell via the ACE2 receptor (Li et al., 2003). ACE2 receptor is significantly expressed in the lung, kidney, blood vessels and also in the mucosa of the oral cavity this could explains the incidence of pneumonia and bronchitis seen in COVID-19 positive patients (Ciaglia et al., 2020). Results of our study show that 6/29 viral encoded proteins (nucleocaspid phosphoproteins, chain b spike glycoprotein, membrane glycoprotein, orf1ab polyprotein, replicase polyprotein 1ab, protein 9b) were significantly increased in COVID-19 positive patients as compared to COVID-19 negative patient. These results show that viral protein quantitation using mass spectrometry could aid in viral diagnosis in COVID-19 patients. Interestingly respiratory specimen of COVID-19 positive patient showed significant increase in the level of ACE 2 receptor. This increase in the ACE2 receptor level in the respiratory specimen could be attributed to a probable increase in ADAM17 activity (Ciaglia et al., 2020) or increase worn out bronchial epithelial cells in the respiratory specimen, though these observation warrants further evaluations.

Deep analysis of the respiratory specimen (complex mixture of biomolecules) not only can aid in the identification of a panel of indicators for SARS-CoV-2 infection but also could provide insight on the pathophysiology of the patients. Thus respiratory specimen could be classified as a liquid biopsy which on detailed analysis could provide clues for diagnostic/ prognostic or therapeutic implication. Keeping this in mind we adopted a multi-omics approach to characterize baseline molecular determinants associated with SARS-CoV-2 diagnosis.

Results of our study clearly segregate COVID-19 positive patients from COVID-19 negative or Influenza A H1N1 pdm 2009 positive patients based on the respiratory specimen proteome. We were able to identify a panel of proteomic/metabolomic or metaproteomic signatures which could be used for screening of SARS-COV2 infection and segregation of COVID-19 positive patients. Panel proposed in the study comprise of proteins linked to interferon activation, viral carcinogenesis, neutrophil and monocyte activation and others, metabolites linked to glycerophospholipid metabolism, histidine metabolism and inositol phosphate metabolism known to be linked to viral infection (Metzner et al., 2008; Thaker et al., 2019).

SARS-CoV-2 infection leads to severe immune activation which is followed by acute respiratory distress (Astuti and Ysrafil, 2020), concordant to the pathogenesis, proteome analysis of the respiratory specimen of COVID-19 positive patients showed significant increase in proteins linked to viral life cycle and carcinogenesis, immune activation including neutrophil, monocyte activation and degranulation, IL-17 signaling, NOD-like receptor signaling, leucocyte transendothelial migration and antigen presentation which are known to be increase with the increase in SARS-CoV-2 infection (Li et al., 2020). Viral infection are also known for metabolic reprograming of host (Thaker et al., 2019) and proteome analysis of the respiratory specimen showed that there is significant increase in proteins associated to glucose metabolism suggesting that SARS-CoV-2 induces energy metabolism (Supplementary Figure 18). Interestingly we found increase in proteins linked to fluid shear stress and bacterial invasion of the epithelial cells in the respiratory specimen. This suggests that COVID-19 positive patients may have secondary bacterial infections and associated comorbidities which need further evaluation. Decrease in the oxygen caring capacity is a hall mark feature of SARS-CoV-2 infection which leads to acute respiratory distress syndrome (ARDS) (Geier and Geier, 2020) and respiratory specimen of COVID-19 positive patients showed concordant decrease in proteins linked to gas transport particularly oxygen transport (heamoglobin binding, haptoglobin binding), hydrogen peroxide and cofactor catabolic/metabolic process. Decrease in the oxygen transport proteins was linked to an associated decrease in wound healing, regulation of body fluid levels and vesicular transport seen in the respiratory specimen of COVID-19 positive patients. These disturbances can lead to uncontrolled production of inflammatory mediators, contributing to a state of persistent injury in COVID-19 positive patients. Together these observations suggest that the respiratory specimen could provide an insight into the pathophysiology (virus mediated hyper immune activation and decrease in oxygen transport and wound healing) of SARS-CoV-2 infection which could be explored for prognosis and therapeutic intervention.

Concordant to the proteomic analysis, results of the metabolome analysis showed that respiratory specimen of COVID-19 patients is enriched for metabolites linked to unsaturated fatty acids and glycerophospholipid metabolism involved in the early development of virus (Schoggins and Randall, 2013). Further there was significant increase in ubiquinone/terpenoid-quinone biosynthesis, aminoacyl-tRNA biosynthesis along with amino acid metabolism. Whereas metabolites linked to vitamin metabolism and steroid metabolism was significantly decreased in COVID-19 respiratory specimen. These results suggest that respiratory specimen of COVID-19 positive patients is rich in oxidative and inflammatory metabolite and has significantly lower levels of anti-inflammatory steroids or vitamins.

Results of the respiratory proteomics show significant increase in bacterial invasion of the epithelial cells. Further bacterial co-infection aggravates viral infection and increase the duration of the disease (Gu and Korteweg, 2007). In addition, alteration of microbiome (gut) is linked to immune activation in SARS-CoV2 infection (Dhar and Mohanty, 2020). As oropharyngeal microbiome is reflective of lung microbiome (Dickson and Huffnagle, 2015) and SARS-COV2 infection could modulate it. Change in the oropharyngeal microbiome could be reflective of altered pathogenesis in COVID-19. This prompted us to perform metaproteome (microbiome) analysis of the respiratory specimen in COVID-19 positive, negative and H1N1 positive patients. Metaproteome analysis of COVID-19 positive respiratory specimen showed significant increase in Bacteroidetes Order II. Incertae sedis, Bacillales, Burkholderiales, Klebsiella pneumonia, Pasteurellales and other bacterial species known to be increased in lung and upper respiratory pathogenesis such as severe asthma, COPD or SARS (Wu and Segal, 2017) and (Gu and Korteweg, 2007). Interestingly, Lactobacillus salivarius known as a potent antialergic and probiotic was significantly reduced in COVID-19 positive patients (Li et al., 2010). Functionality assessment based on the respiratory metaproteome showed that proteobacteria: enzyme encode pathways such as fatty acid biosynthesis, glycerolipid metabolism, amino acid, energy metabolism and oxidative phosphorylation and firmicutes or tenericute: enzyme encodes pathways aminoacyl-trna biosynthesis and terpenoid backbone biosynthesis show similarity with the respiratory metabolome enriched pathways suggesting to the involvement of bacteria in the metabolic regulation of SARS-CoV-2 infection. Together these results suggest that SARS-CoV-2 infection modulates the respiratory microbiome and functionality thus providing destabilized microenvironment which may predispose patients to respiratory distress such as allergic cough, fever and others.

Viral infection induces angiogenesis (Cerimele et al., 2003) and is known to increase proteins processing/maturation and glycosylphosphatidylinositol (GPI) anchor biosynthesis (Metzner et al., 2008). Results of the global cross correlation analysis were concordant and showed that the viral proteome share a direct correlation with bacterial metaproteome and proteins or metabolites linked to viral life cycle, regulation of angiogenesis, vasculature development, protein processing/maturation, beta-alanine metabolism, pantothenate and CoA biosynthesis and glycosylphosphatidylinositol (GPI) anchor biosynthesis. Results of the multi-omics integration also show that with the increase in SARS-CoV-2 viraemia there is significant increase in energy metabolism (Pentose phosphate pathway, Glycolysis), Glycerophospholipid metabolism, Purine metabolism and inflammation (IL-17 signaling pathway) in COVID-19 positive patients these observation correlates with classical features of virus replication, metabolic regulation and inflammation (Thaker et al., 2019). Recently, Shen et.al (Shen et al., 2020) documented dysregulated lipid metabolism, increase in acute phase reactant, immune activation (macrophage, platelets) in severe COVID-19 patients. Our results was able to recapitulate the findings and showed that respiratory specimen of COVID-19 positive patients have significant increase in modules associated to immune activation generic cluster, enriched in monocytes and platelet activation module I and II. We additionally showed that these modules show direct correlation with the metabolites linked to arginine biosynthesis, cysteine and methionine metabolism and Aminoacyl-tRNA biosynthesis. Arginine metabolism is a classical feature of inflammatory macrophages and increase in arginine biosynthesis in monocytes suggests inflammatory activation of monocytes (Rath et al., 2014). Interestingly, multi-omics analysis was also able to segregated COVID-19 asymptomatic from the symptomatic. Molecules such as AKR1A1 (Aldo-keto reductase family 1 member A), COPE (Coatomer subunit epsilon) and C02571 (L-Acetylcarnitine) were asymptomatic specific while symptomatic COVID-19 patients showed significant increase in Klebsiella pneumoniae, CTBP2 (C-terminal-binding protein 2) and 4-alpha-Methyl-5-alpha-cholest-7-en-3-one suggesting that the multi-omics profile could also identify asymptomatic (silent carrier of infection) which could mediate an exponential increase in the infection rate.

Finally, of the panel of molecular signatures (Table 1) capable of segregating COVID-19 positive patients. We validated MX1 (MX Dynamin like GTPase 1) and WARS (Tryptophan-- tRNA ligase) in 200 COVID-19 suspects. MX1 is an effector anti-viral protein which modulate the type-I interferon mediated inflammatory response in lungs (Makris et al., 2017). MX1 is activated in response to novel viruses for which the body has no immune defense (antibodies) (Haller et al., 2015). SARS-COV2 as a novel virus mediates cellular entry via ACE2 receptor or via membrane fusion (cleavage of the spike glycoprotein by TMPRSS2 (type II transmembrane serine protease)). This increases expression of both MX1 and TMPRSS2 via interferon regulatory factor 1 (IRF-1) (Panda et al., 2019). Increase in MX1 (chemo attractant) mediate infiltration of immune cells (Neutrophil, Monocyte) and induces interferon response (Haller et al., 2015). Increase in site specific interferon response increases secretion of WARS (housekeeping enzyme) which activates monocytes to macrophage via TLR2-TLR4 pathway (Jin, 2019) followed by other immune cell activation which may lead to cytokine storm seen in COVID-19 patients. Thus, over expression of MX1 and WARS in response to SARS-COV2 infection in the respiratory specimen serves as attractive molecular targets and were cherry picked for evaluation of diagnostic potential in 200 COVID-19 suspects. Results of our study showed that MX1 >30pg/ml and WARS >25ng/ml in the respiratory specimen was able to segregate COVID-19 positive from COVID-19 negative patients with a combined diagnostic efficiency of 94% and a predictive accuracy of 86%. These results potentiate the utility of MX1 and WARS in segregation of COVID-19 patients.

Due to biosafety constrains in SARS-CoV-2 infection large number of samples were not collected nor analyzed in the study. Only previously archived (left over) respiratory specimen of the patients coming for COVID-19 testing at the Institute of Liver and Biliary Sciences New Delhi, were analyzed in the study. Detailed demographic profile and severity indices of the patients could not be collected as only archived samples were used for analysis. Parallel reaction monitoring (PRM) based assessment of viral proteins could help in absolute quantitation of viral proteins but was not performed in the study. We performed absolute quantitation of two best indicators of COVID-19 positivity. However, molecular targets such as metabolites, metaproteins and viral proteins amongst the diagnostic panel for COVID-19 segregation warrant further validation.

In conclusion, our study presents a multi-omics investigation of respiratory specimen from COVID-19 positive and controls. Our data provides a systemic overview of the host response induced by SARS-COV2 infection in the respiratory specimen. MX1 (MX Dynamin like GTPase 1) and WARS (Tryptophan--tRNA ligase) efficiently diagnosed COVID-19 positive patients and could be used for mass screening of COVID-19 patients. Systemic overview of the respiratory specimen could provide useful prognostic and therapeutic indications in the ongoing battle against the COVID-19 pandemic.

### STAR★Methods

**STAR★Methods:**
| REAGENT or RESOURCE | SOURCE | IDENTIFIER |
| --- | --- | --- |
| **Biological Samples** |  |  |
| *Discovery Cohort:*<br>Serum samples from 20 COVID-19 positive patients and 20 COVID-19 negative patients and 05 H1N1 subjects in discovery cohort.<br>*Validation:* Targets were validated on 200 COVID-19 patients. | ILBS, Hospital | (Supplementary Table 7) in this Manuscript. |
| **Chemicals, Peptides, and Recombinant Proteins** |  |  |
| Triethylammonium bicarbonate buffer (TEAB) | Thermo Fisher Scientific | Cat # PI90114 |
| Urea | Thermo Fisher Scientific | Cat # 29700 |
| Tris (2-carboxyethyl) phosphine (TCEP) | Thermo Fisher Scientific | Cat # T2556 |
| Iodoacetamide (IAA) | Sigma-Aldrich | Cat # I6125 |
| Dithiothreitol | Sigma-Aldrich | Cat # D0632 |
| Trypsin | Promega | Cat # V5111 |
| Acetone | JT Baker | Cat # 9006-03 |
| Water | Thermo Fisher Scientific | Cat # W6-4 |
| Acetonitrile | JT Baker | Cat # 14650359 |
| Formic acid (FA) | Sigma-Aldrich | Cat # F0507 |
| Ammonium hydroxide solution | Sigma-Aldrich | Cat # 221228 |
| Methanol | Sigma-Aldrich | Cat # 34860 |
| C18 Spin Columns | Thermo Fisher Scientific | Cat # 89870 |
| Bradford Total protein assay kit | Thermo Fisher Scientific | Cat # 22662 |
| **ELISA / Antibodies:** |  |  |
| MX1 ELISA | Cusabio | Cat: No; csb-El015249HU |
| WARS ELISA | Elabscience | Cat: No; E-EL-H1874 |
| **Software and Algorithms** |  |  |
| Xcalibur | Thermo Fisher Scientific | Cat # OPTON-30965 |
| Proteome Discoverer Version 2.4.1.15 | Thermo Fisher Scientific | https://www.thermofisher.com/us/en/home/industrial/mass-spectrometry/liquidchromatography-mass-spectrometry-lc-ms/lc-ms-software/multi-omics-data-analysis/proteome-discoverer-software.html. |
| Compound Discoverer Software 3.1 | Thermo Fisher Scientific | https://www.thermofisher.com/order/catalog/product/OP TON-30925#/OPTON-30925 |
| Metaboanalyst 4.0 | webserver | https://www.metaboanalyst.ca/MetaboAnalyst/home.xhtml |
| R version 3.5.2 | R Project | https://www.r-project.org |
| Cytoscape | United States (Java) | https://cytoscape.org/ |
| unipept | webserver | https://unipept.ugent.be/ |

#### Resource Availability

##### Lead Contact

Further information and requests for resources and reagents should be directed to and will be fulfilled by the Lead Contact, Dr. Jaswinder Singh Maras and Dr. Shiv Kumar Sarin.

Raw Data: The raw data for the manuscript is available on request to the lead contact Dr. Jaswinder Singh Maras and Prof. Shiv Kumar Sarin.

##### Sample/Patient selection

A cross sectional study was planned, and previously archived (left over) respiratory specimens (combined oropharyngeal and nasopharyngeal swabs in viral transport media) of the patients coming for COVID-19 testing at the Institute of Liver and Biliary Sciences New Delhi, were selected for the study. Multi-omics analysis was performed on respiratory specimen of RT-PCR proven COVID-19 positive, negative (n=20) or H1N1 patients. Fourteen of the 20 COVID-19 positive patients showed classical symptoms including fever, sore throat, cough and breathlessness, whereas 6 patients were asymptomatic yet positive for COVID-19 and were included in the study.

##### Discovery

For the discovery phase 150 ul of the respiratory specimen from COVID-19 positive patients (n=20), COVID-19 negative patients (n=20) and respiratory diseases control Influenza A H1N1 pdm 2009 positive samples (n=5) were subjected to proteomics (SARS-CoV-2 virus proteins and linked proteins and host proteins), metaproteomics (bacterial peptide) and metabolomics evaluations (Supplementary Table 7). Baseline demographic profiles were recorded and baseline samples were used for multiomics evaluation and correlation to disease pathogenesis. In brief, for proteomics and metaproteomics application, 4% SDS was added to 100 ul of the respiratory specimen, kept at 95LJ for 15 min (viral deactivation). Total proteins in the respiratory specimen were precipitated by adding 6X cold acetone followed by centrifugation at 13000 rpm for 10 mins. The precipitated proteins were re-dissolved in ammonium bicarbonate buffer (pH-7) and were subjected to LC MS/MS evaluation of virome, host proteome and metaproteome detailed below. The left over 50 ul of the respiratory specimen was subjected to methanol precipitation followed by total metabolome evaluation as detailed below.

##### Validation

An annotated set of 200 COVID-19 suspect patients (100 COVID-19 positive and 100 COVID-19 negative; Supplementary Table 7) was taken to validate the most significant indicators of COVID-19 diagnosis in the discovery phase. The sensitivity/specificity and predictive accuracy was analyzed.

##### Proteomics of the respiratory specimen

Total proteins were isolated from the respiratory specimen of the study groups. 100 ug equivalent protein was subjected to reduction, alkylation followed by digestion for 16-20 h at 37°C using sequencing-grade modified trypsin: proteins (1ug:20ug w/w). The samples were desalted and subjected to LC-MS/MS analysis. The peptides were eluted by a 3–95% gradient of buffer B (aqueous 80% acetonitrile in 0.1% formic acid) at a flow rate of 300 nL/min for about 60 minutes on a 25-cm analytical C18 column (C18, 3 μm, 100 Å) The peptides were ionized by nano-electrospray and subsequent tandem mass spectrometry (MS/MS) on a Q-ExactiveTM Plus (Thermo Fisher Scientific, San Jose, CA, United States). The peptides were analyzed using a mass spectrometer with the collision-induced dissociation mode with the electrospray voltage was 2.3 kV. Analysis on the orbitrap was performed with full scan MS spectra with a resolution of 70,000 from m/z 350 to 1800.The MS/MS data was analyzed by Proteome Discoverer (version 2.0, Thermo Fisher Scientific, Waltham, MA, United States) using the uniprot COVID-19 database (https://covid-19.uniprot.org) containing 14 SARS2, 15 CVHSA and 12 human protein for virome and virus linked proteins detection. The MS/MS data was reanalyzed by Proteome Discoverer (version 2.0, Thermo Fisher Scientific, Waltham, MA, United States) using the uniprot homo sapiens (Human) database (UP000005640) and human proteome with Mascot algorithm (Mascot 2.4, Matrix Science). Significant proteins were identified at (p<0.05) and q values (p<0.05). The threshold of false discovery rate was kept at 0.01. The identified proteins were subjected to standard statistical analysis and network and pathway analysis (Malehmir et al., 2019; Zhou et al., 2019).

##### Metabolomics of the respiratory specimen

Metabolites were extracted from the 50ul of respiratory specimen using organic phase extraction protocol (Song et al., 2017). To 50 ul of respiratory specimen, 500 ul methanol was added and kept overnight at −20 degree for protein precipitation. The samples were centrifuged at 13.3*1000 rpm for 10 min and the supernatant was dried under vacuum. The dried samples were reconstituted with (5:95:5) 5% acetonitrile: 95% water: 5% internal standards at known concentrations, for reverse-phase chromatography by using C18 column (Thermo Scientific(tm)25003102130: 3 µm, 2.1 mm, 100 mm) using ultra-high performance liquid chromatographic system followed by high-resolution mass spectrometry (MS) (Starkel et al., 2018). Metabolite features were extracted using compound discoverer 3.0 (Thermo) (Basu et al., 2017). Annotation of the features was performed using mass list searches (Boudah et al., 2014; Starkel et al., 2018), mzCloud™ (www.mzcloud.org) and mummichog (Song et al., 2017). Identified and annotated features were subjected to log normalization and pareto-scaling using metaboanalyst 4.0 (http://metaboanalyst.ca.) server (Chong et al., 2018) and into SIMCA P12 software (Umetrix, Sweden) for multivariate projection analyses, such as principal component analysis and partial least square discriminant analysis. Pathway enrichment patterns were analyzed using Metaboanalyst 4.0 (Chong et al., 2018).

##### Metaproteome (Microbiome) analysis of the respiratory specimens

Proteins were isolated from the respiratory specimen of the study groups. The isolated proteins were reduced, alkylated and digested using trypsin followed by mass spectrometry analysis similar to that stated in respiratory specimen proteome analysis section. The MS/MS data was acquired and analyzed by Proteome Discoverer (version 2.0, Thermo Fisher Scientific, Waltham, MA, United States) using the bacterial/fungal sequence (UniprotSwP_20170609, with sequences 467231 and MG_BG_UPSP with sequences 2019194). This was cross validated using Mascot algorithm (Mascot 2.4, Matrix Science) specifically for all possible microbial species. In brief, significant peptide groups were identified at (p<0.05) and q values (p<0.05) and the false discovery rate at 0.01. Only rank-1 peptides with Peptide Sequence Match (PSM)>3 were subjected to biodiversity and functional analysis using unipept (Zhang et al., 2018). Peptides mapping to eukaryotic, fungal and viral database were rejected and only bacterial species associated peptides were segregated and were subjected to statistical, functional and biodiversity analysis (Zhang et al., 2018).

##### Global cross-correlation, clustering and integration analysis

Differentially expressed viral proteome (DEVP), differentially expressed host proteome (DEPs), differentially expressed metabolites (DEMs) and differentially expressed metaproteins (DEMP) were identified and subjected to cross correlation and clustering analysis (r2>0.5,p<0.05). Significant and prominent clusters were identified and subjected to pathway analysis; for proteins (enricher/KEGG) and metabolites (KEGG/metaboanalyst). This was followed by developing a global cross-correlation map between the virome, metaproteome and the pathways linked to the proteins and metabolites using cytoscape (https://cytoscape.org/) (Shannon et al., 2003).

ELISA quantitation: MX1 (Cat: No; csb-El015249HU) with a detection limit of 15.6 pg/mL-1000 pg/mL, sensitivity 3.9 pg/mL and WARS (Cat: No; E-EL-H1874) with a detection range 0.78—50 ng/mL and sensitivity of 0.47 ng/mL were used for validation as per manufacture protocol.

#### Statistical Analyses

Results are shown as mean and standard deviation unless indicated otherwise. Statically analysis was performed using Graph Pad Prism v6, SPSS V20 and P-values of < 0.05using Benjamini & Hochberg correction was considered statistically significant. Unpaired (two-tail) Student’s t test, Mann-Whitney U test were performed for comparison of two groups. For comparison among more than two groups, one-way analysis of variance, Kruskal-Wallis test was performed. All correlations were performed using Spearman correlation analysis and R^2^>0.5, p<0.05 was considered as statically significant. For the multi-omics analysis, features with over 80% missing values in particular group were neglected further feature with less than 80% missing values in particular group were imputed with the minimal value of the feature in the assigned group. Data for virome, proteome, metabolome and metaproteome analysis was log normalized and subjected to Perato scaling this was followed by calculation of Log2 Fold change for each pair of comparing group. Differentially expressed virome, proteome, metabolome or metaproteome were identified at p<0.05 and fold change >±1.5 FC. From the discovery cohort important viral proteins, host proteins, metabolites and metaproteins were selected for validation based on random forest analysis and area under the curve (AUROC) analysis (AUROC>0.8,p<0.05). Prediction class probability and predictive accuracy (for most important variables) was calculated in the discovery cohort and cross validated in the validation cohort.

## Data Availability

Raw Data: The raw data for the manuscript is available on request to the lead contact Dr. Jaswinder Singh Maras

## Abbreviation

COVID-19: Corona virus diseases 2019
SARS-CoV-2: severe acute respiratory syndrome coronavirus 2

## Supplementary Figures

**Supplementary Figure 1:**
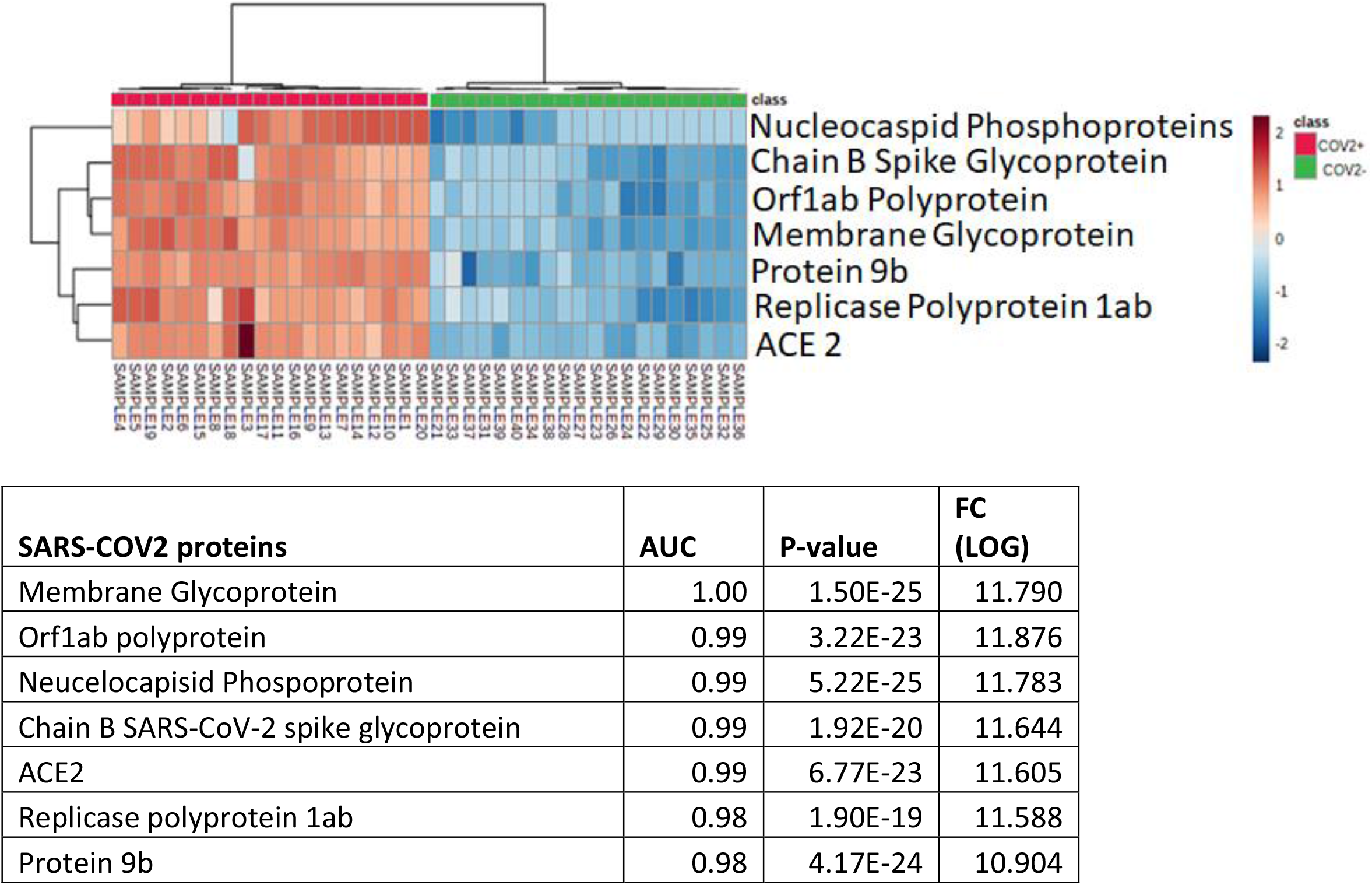
Heat map and hierarchical cluster analysis for the viral proteins identified (p<0.05) show clear segregation of COVID-19 positive (Red bar) from COVID-19 negative (Green bar) patients. The expression is given in the range as Dark brown= upregulated, Blue= downregulated and white= unregulated. AUROC analysis shows clear distinction of SARS-COV2 proteins in Positive patients vs. negative patients.

**Supplementary Figure 2:**
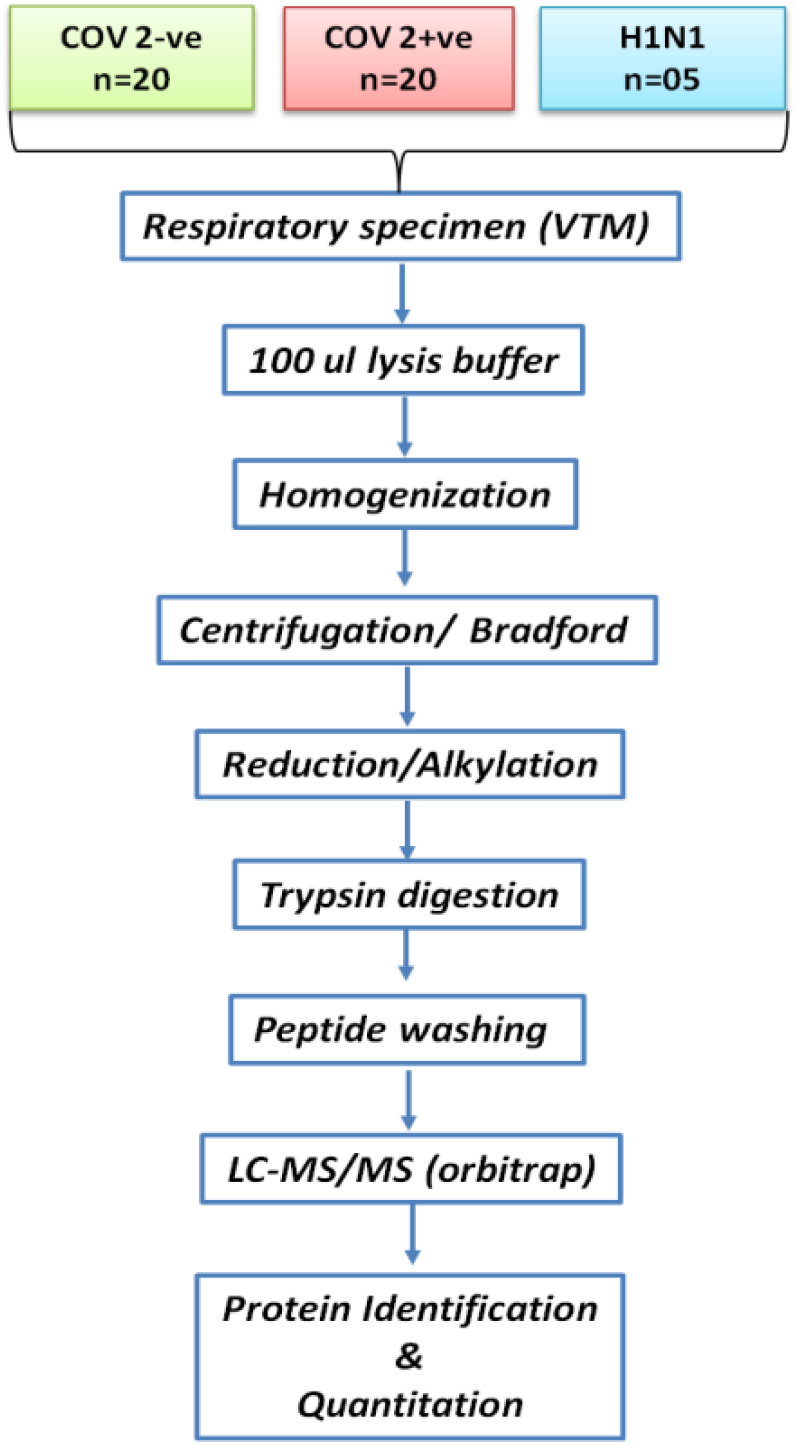
Schema for proteomic sample preparation used in the study.

**Supplementary Figure 3:**
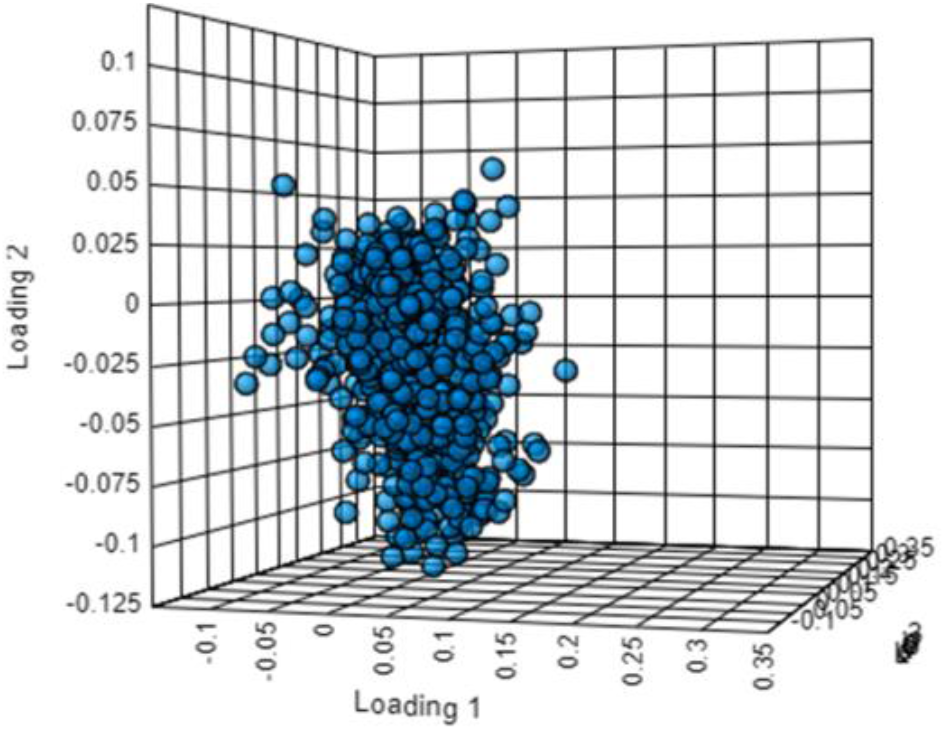
Loading plot for the partial least square discriminant analysis showing clear segregation of COVID-19 positive patients from COVID-19 negative and Influenza A H1N1 pdm 2009 positive patients based on proteomic evaluation.

**Supplementary Figure 4:**
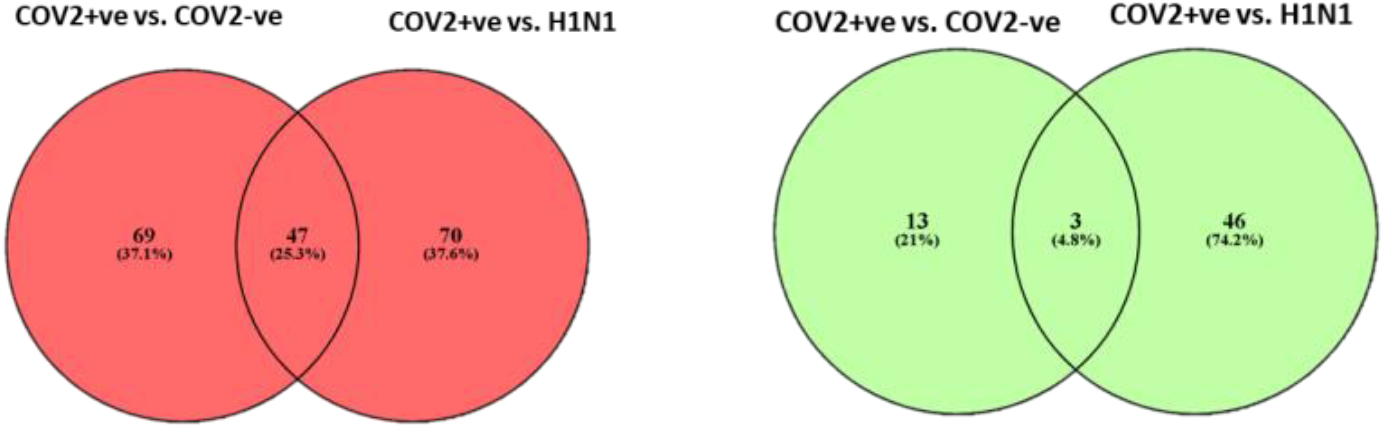
Total no of proteins differentially regulated and their intersection in COV2+ vs. COV2- and COV2+ vs. Influenza A H1N1 pdm 2009 positive cases is shown. Red denotes proteins up regulated and green denoted the proteins down regulated.

**Supplementary Figure 5:**
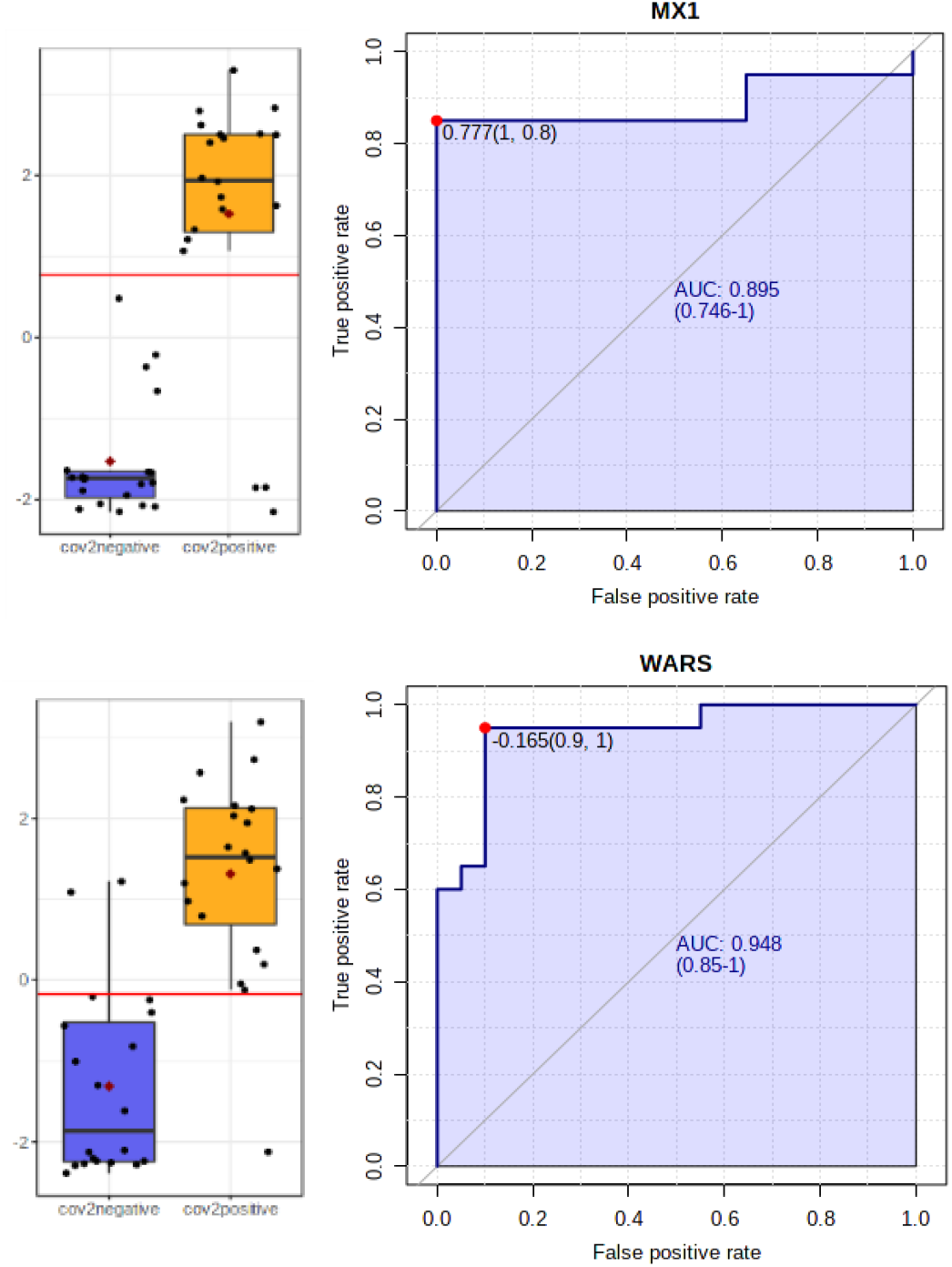
Univariate area under the receiver operating curve analysis for MX1 (AUC=0.895 CI (0.75-1)) and WARS (AUC=0.948 CI (0.85-1))

**Supplementary Figure 6:**
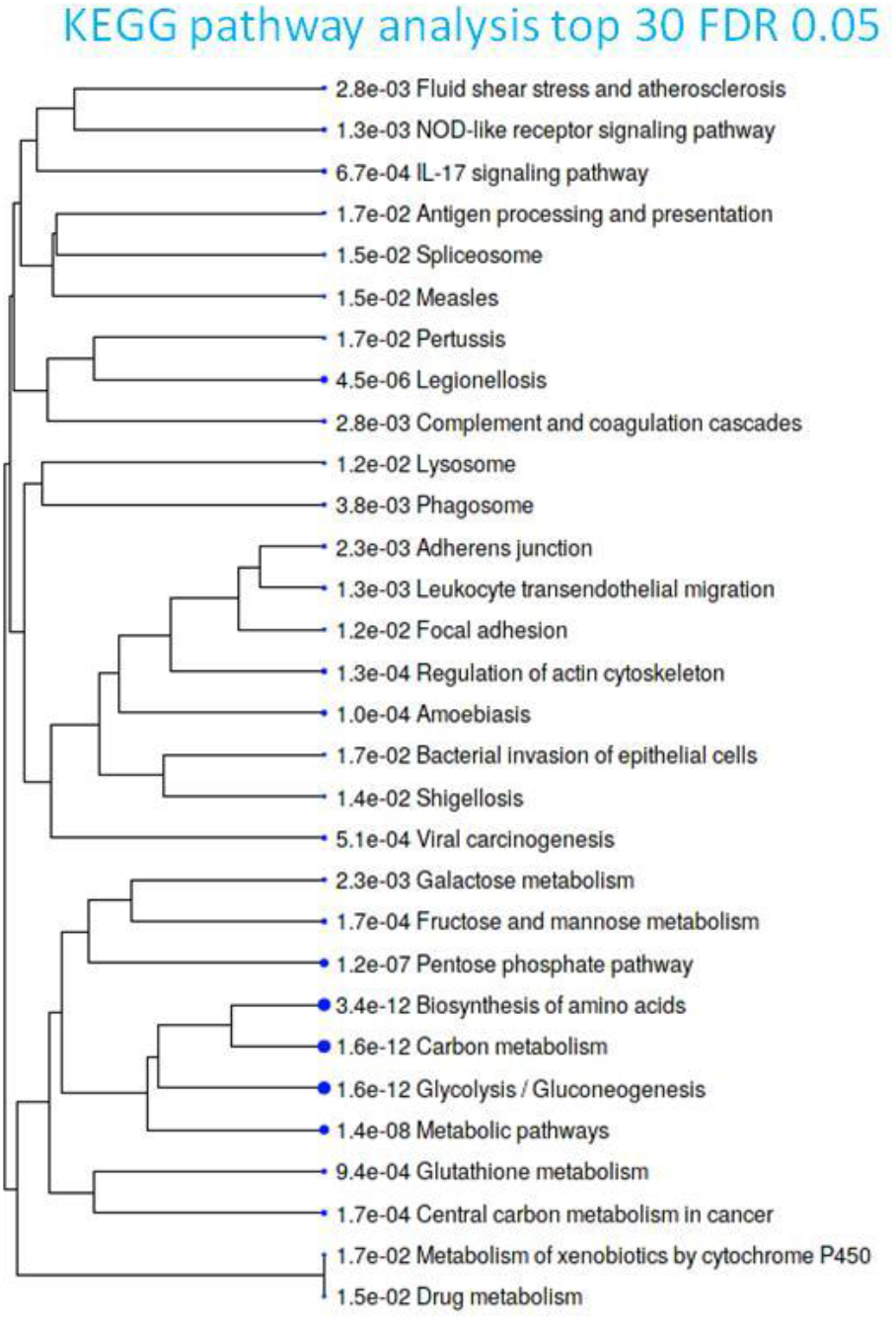
A hierarchical clustering tree summarizing the correlation among significant pathways upregulated in COVID-19 positive respiratory specimen compared to COVID-19 negative or Influenza A H1N1 pdm 2009 positive cases. Pathways with many shared genes are clustered together. Bigger dots indicate more significant P-values.

**Supplementary Figure 7:**
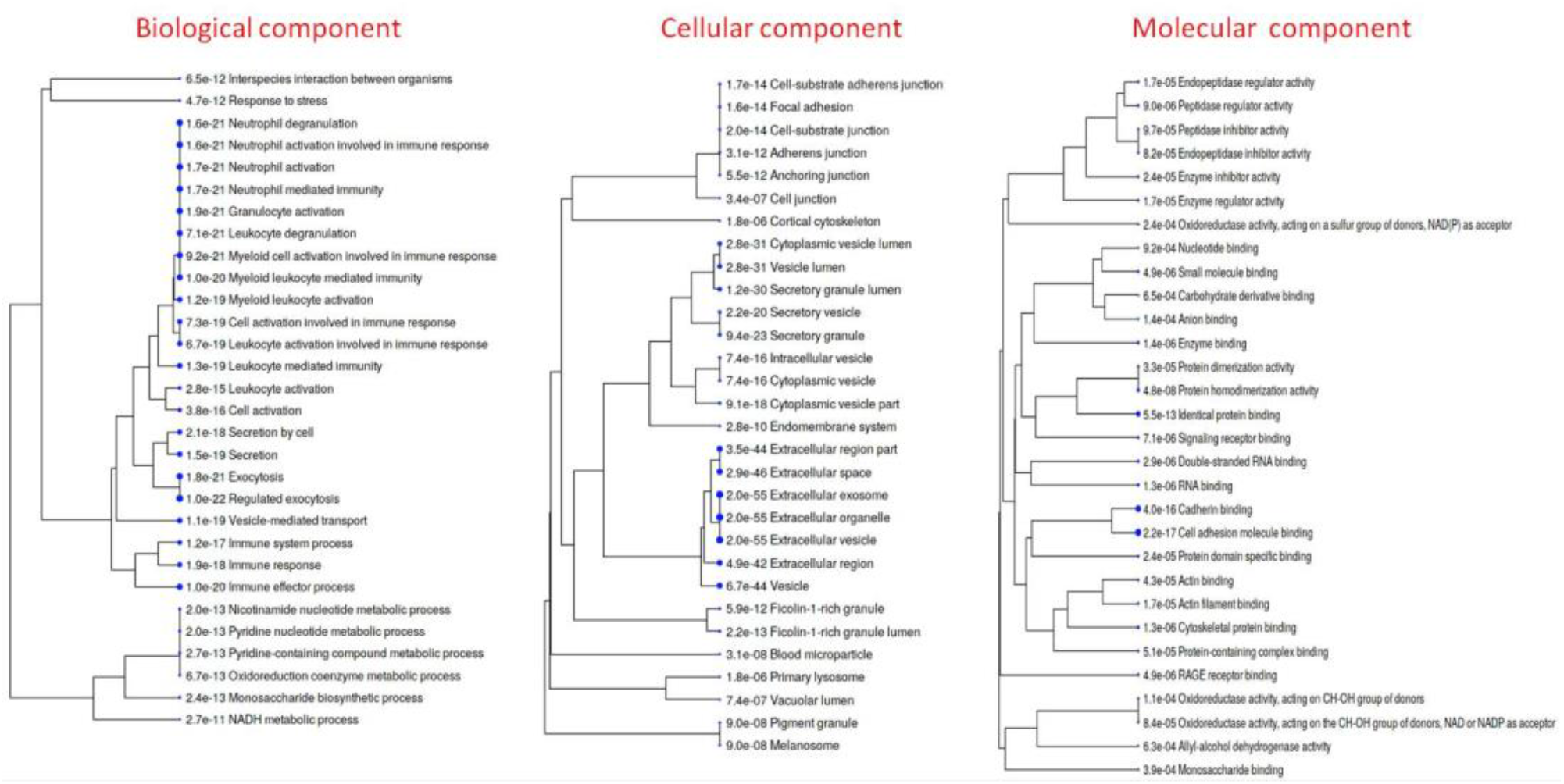
A hierarchical clustering tree summarizing the correlation among significant pathways upregulated in COVID-19 positive respiratory specimen compared to COVID-19 negative or Influenza A H1N1 pdm 2009 positive cases. Pathways with many shared genes are clustered together. Bigger dots indicate more significant P-values.

**Supplementary Figure 8:**
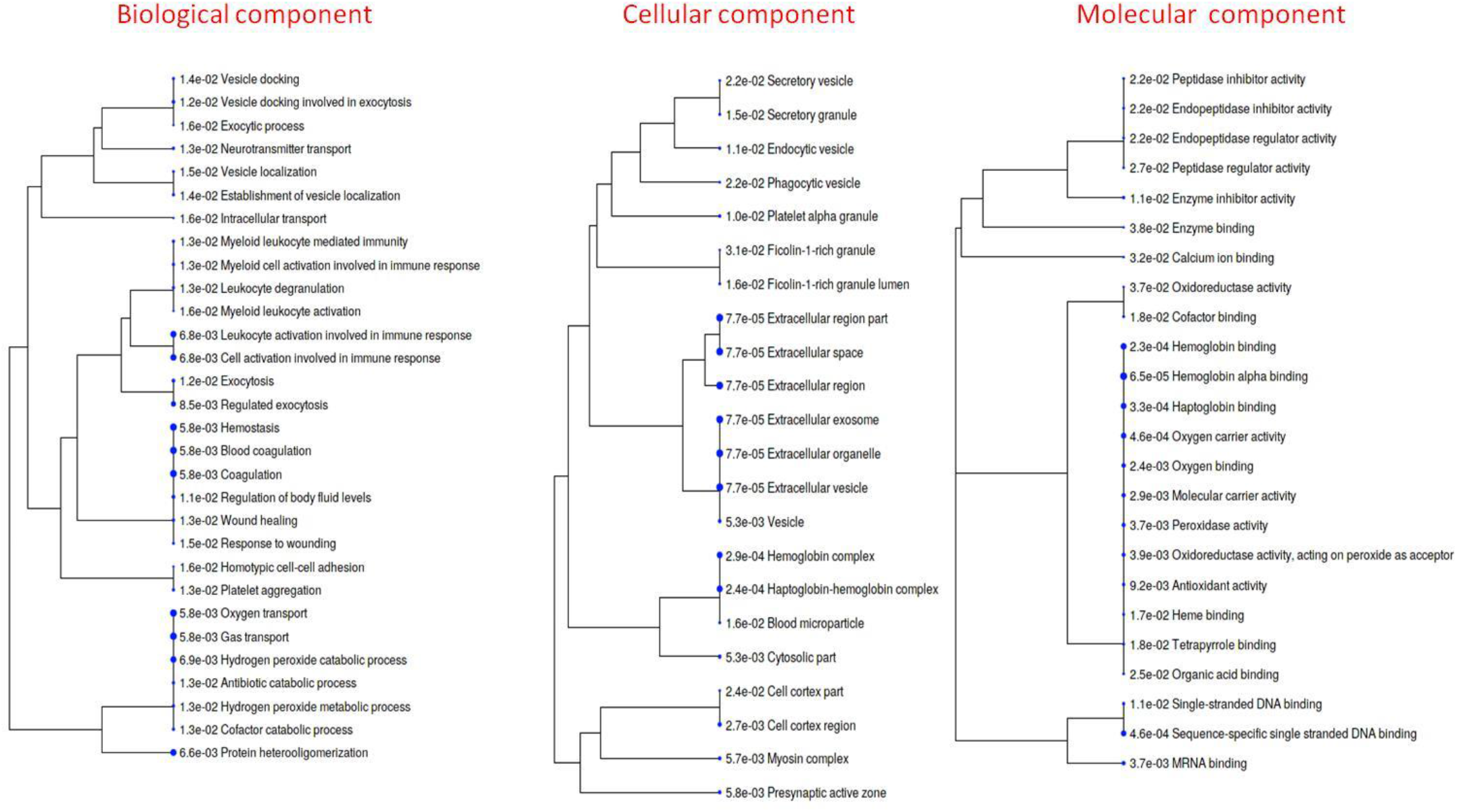
A hierarchical clustering tree summarizing the correlation among significant pathways downregulated in COVID-19 positive respiratory specimen compared to COVID-19 negative or Influenza A H1N1 pdm 2009 positive cases. Pathways with many shared genes are clustered together. Bigger dots indicate more significant P-values.

**Supplementary Figure 9:**
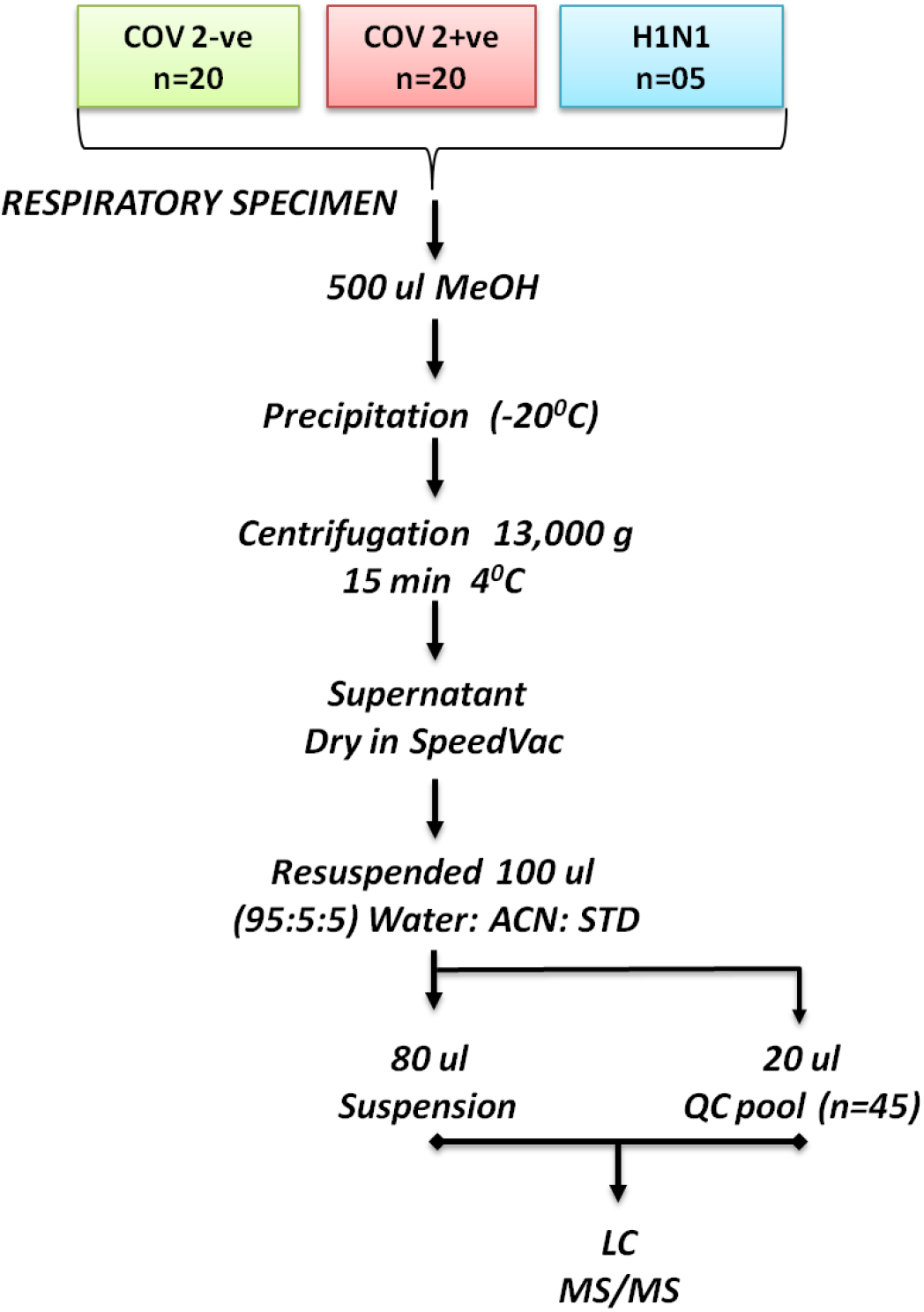
Schema for metabolomics analysis used in the study.

**Supplementary Figure 10:**
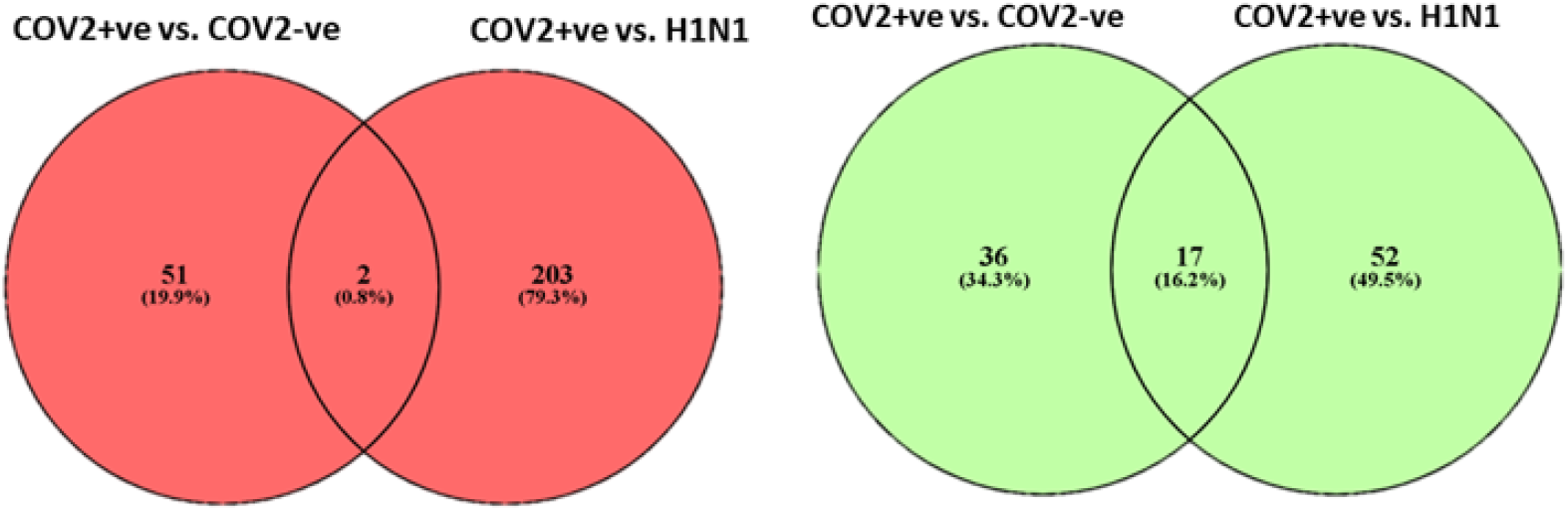
Total no of metabolites differentially regulated and their intersection in COV2+ vs. COV2- and COV2+ vs. Influenza A H1N1 pdm 2009 positive cases is shown. Red denotes proteins up regulated and green denoted the proteins down regulated.

**Supplementary Figure 11:**
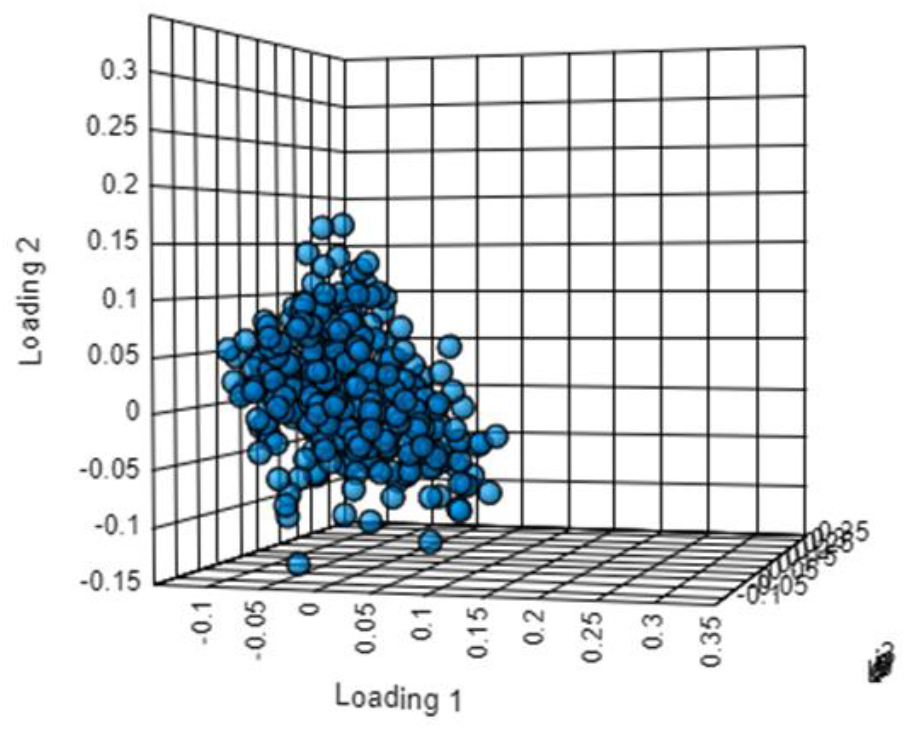
Loading plot for the partial least square discriminant analysis showing clear segregation of COVID-19 positive patients from COVID-19 negative and Influenza A H1N1 pdm 2009 positive cases patients based on metabolomics evaluation.

**Supplementary Figure 12:**
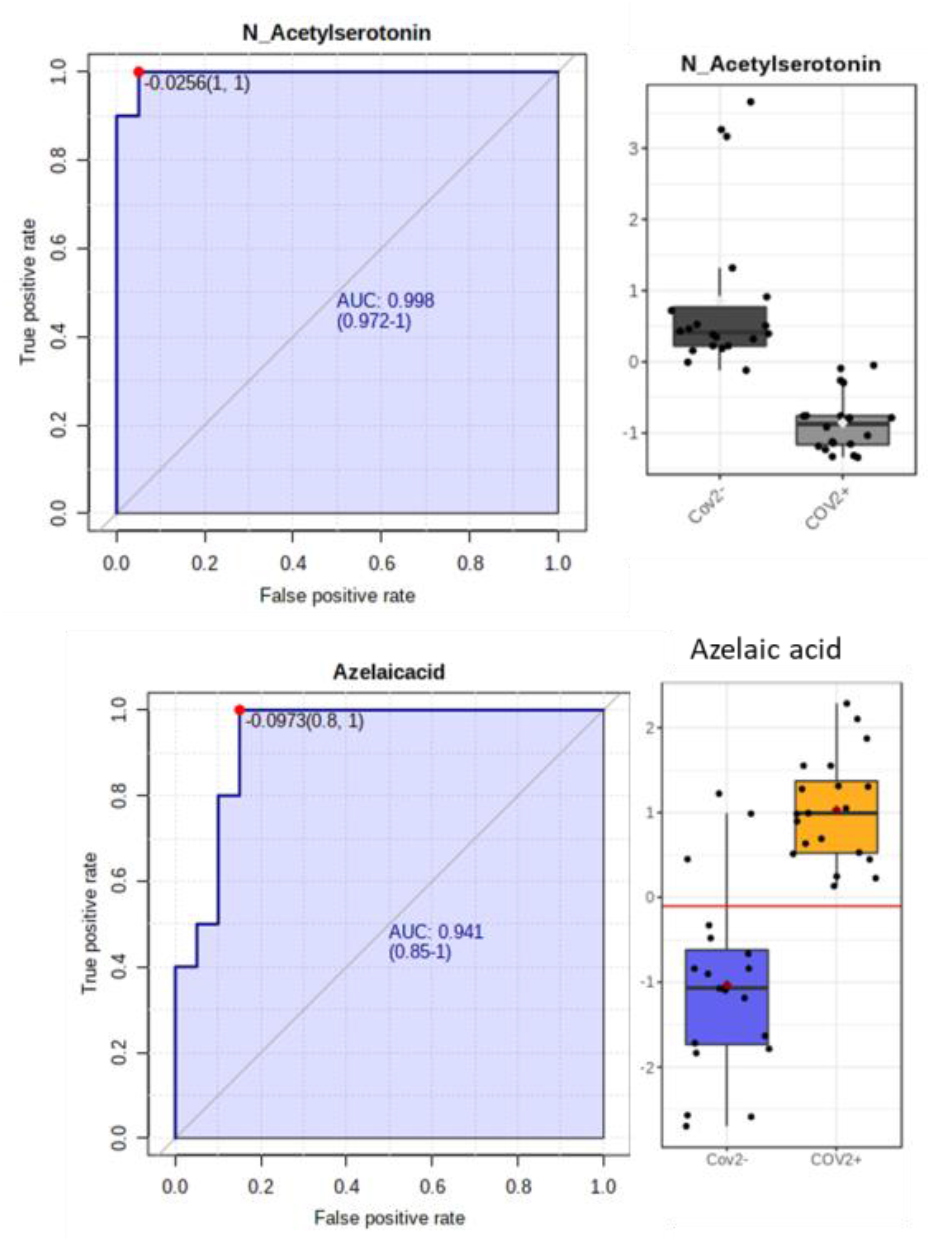
Univariate area under the receiver operating curve analysis for NAcetylserotinin(AUC=0.99 CI (0.975-1)) and Azelaic acid (AUC=0.941 CI (0.85-1))

**Supplementary Figure 13:**
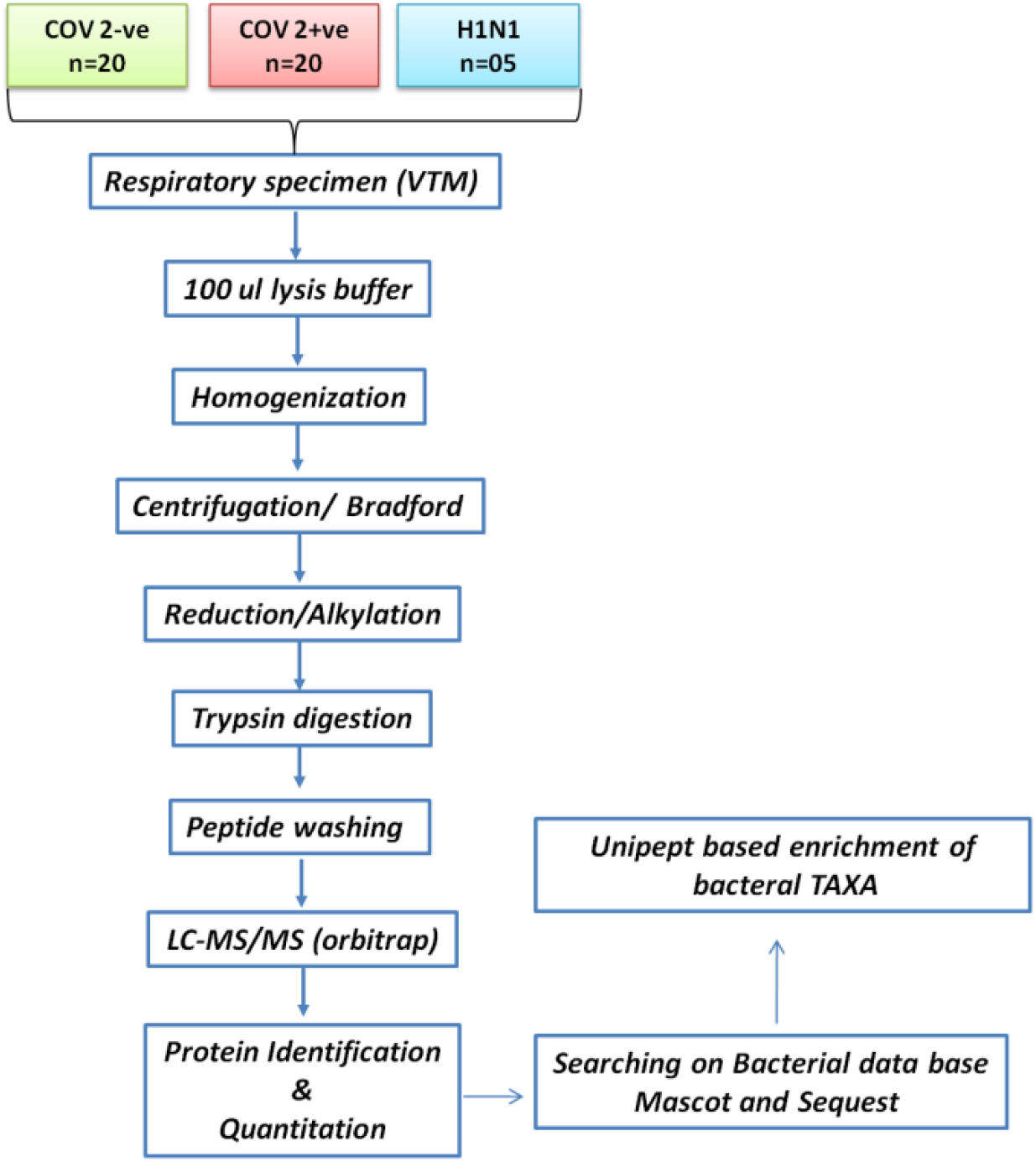
Schema for metaproteomics analysis used in the study.

**Supplementary Figure 14:**
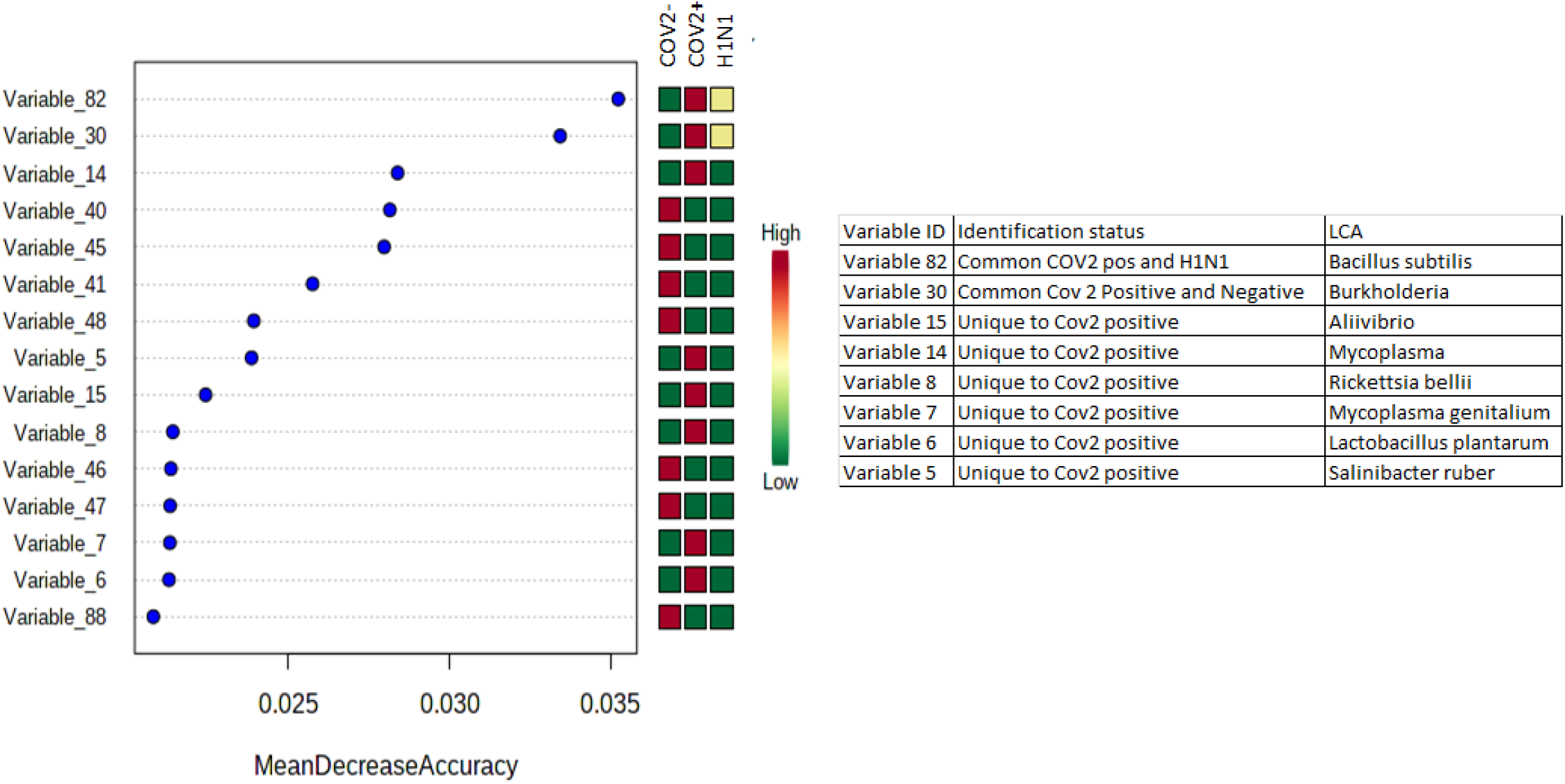
Random forest analysis and mean decrease in accuracy plot showing the mean decrease in accuracy of the metaproteome along with their expression status Red = upregulated and Green= downregulated and yellow= unchanged in COVID-19 positive as compared to COVID-19 negative or Influenza A H1N1 pdm 2009 positive cases patients.

**Supplementary Figure 15:**
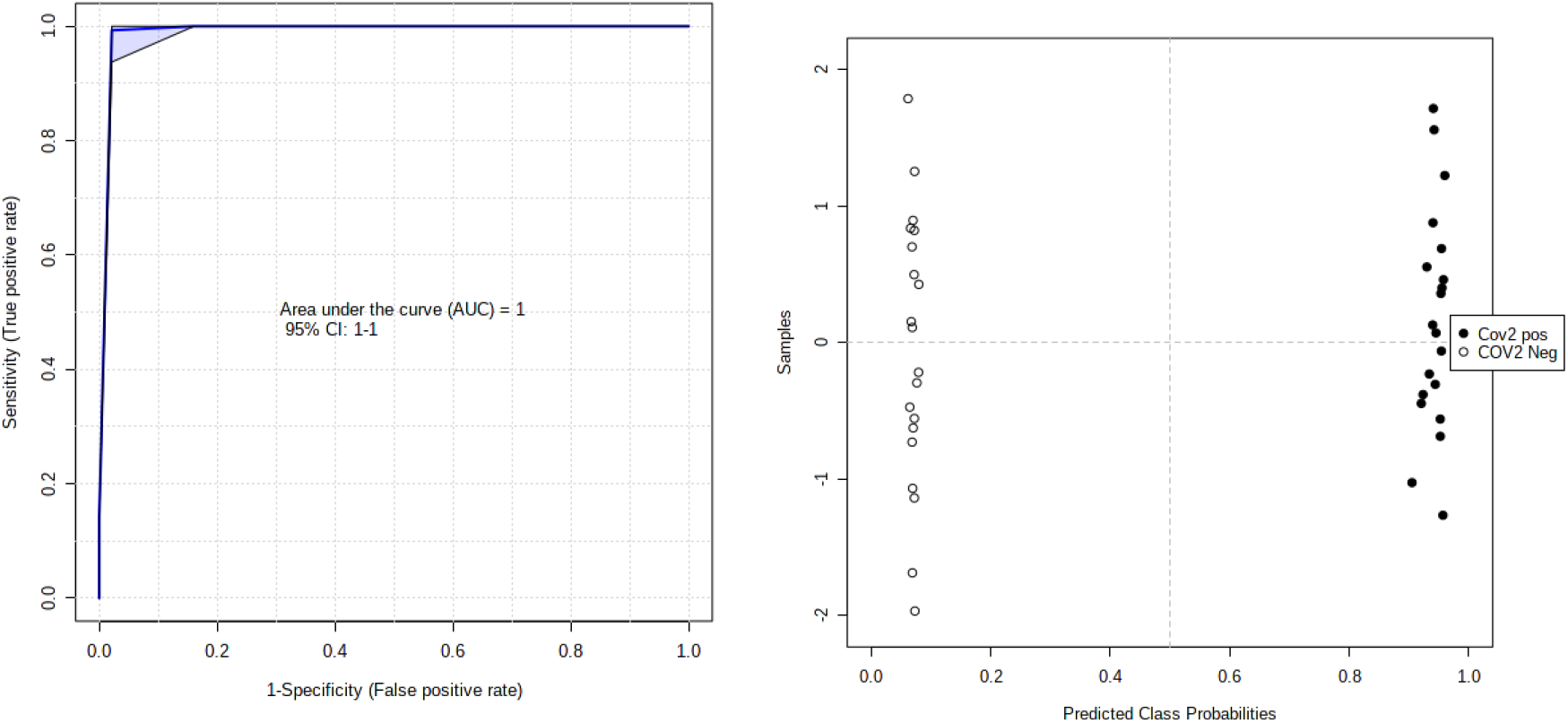
Area under the receiver operating curve analysis for Variable 80 and Variable 30 together show AUC=1 CI (1-1) p<0.05 and Prediction class probability with a predictive accuracy of 100%.

**Supplementary Figure 16:**
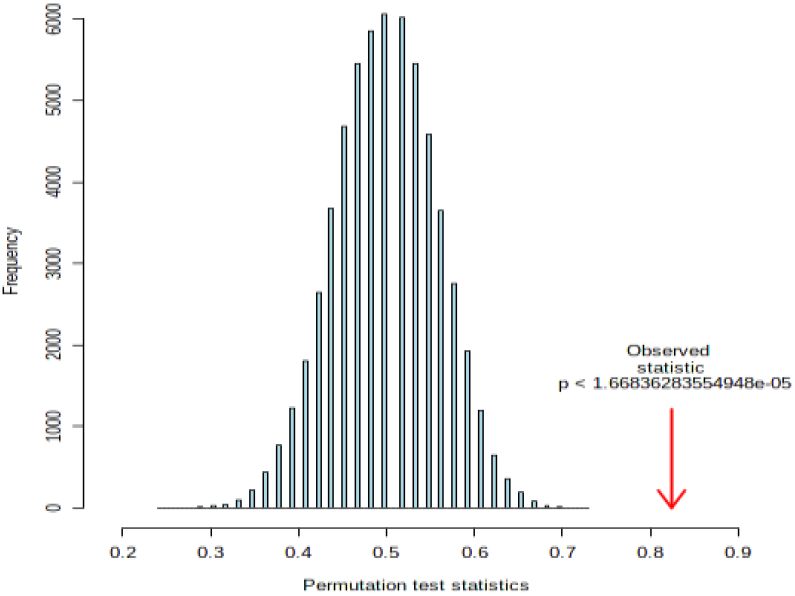
Permutation statistical analysis for prediction class probability of MX1 (1000 permutation) show p<0.01.

**Supplementary Figure 17:**
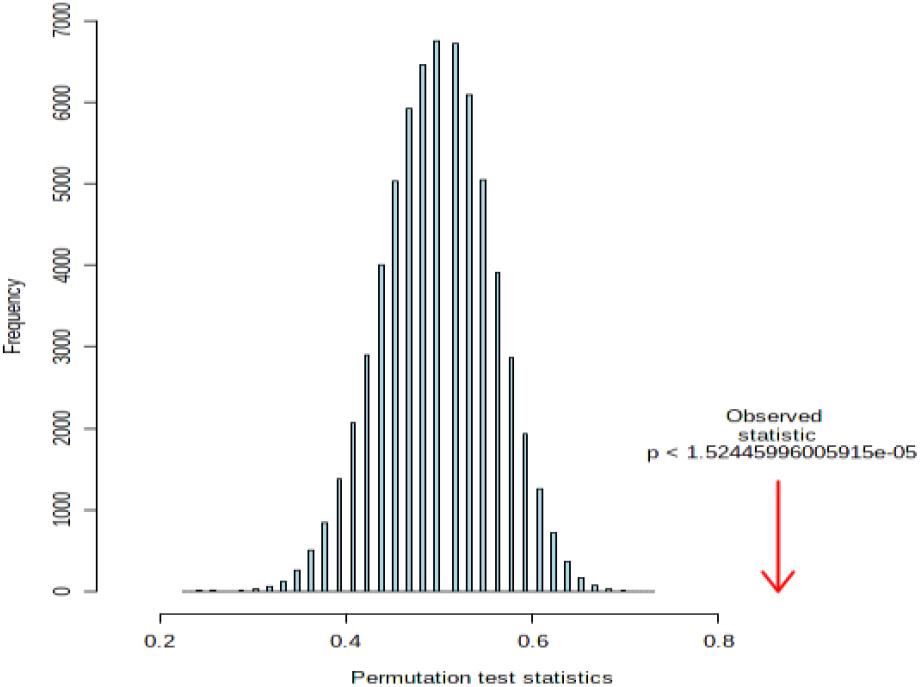
Permutation statistical analysis for prediction class probability of MX1 and WARS (1000 permutation) together show p<0.01.

**Supplementary Figure 18:**
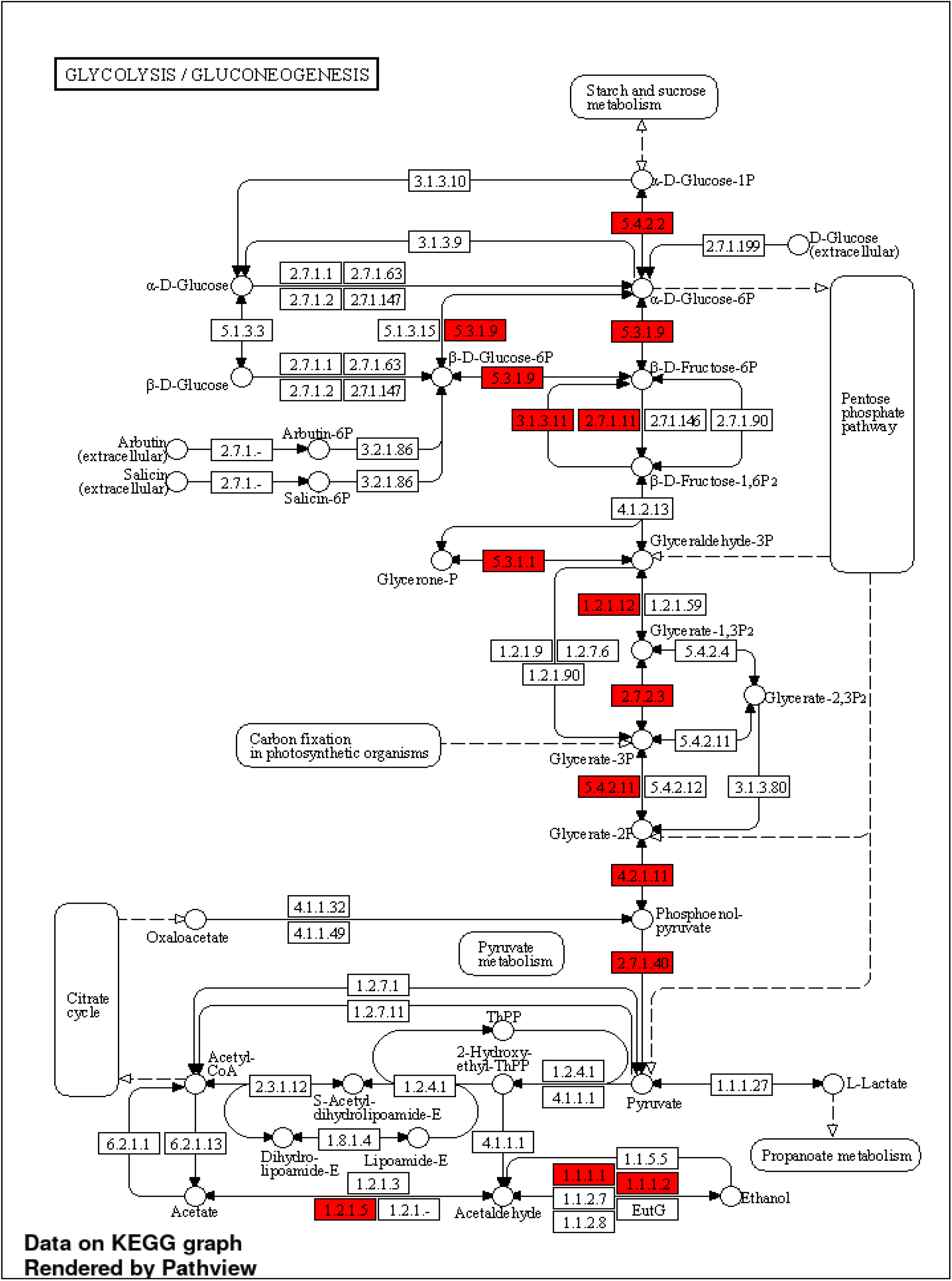
KEGG pathway enrichment analysis for proteins upregulated in the respiratory specimen of COVID-19 positive patients as compared to COVID-19 negative patients showing significant enrichment of proteins lined to Glycolysis/ Gluconeogenesis.

## Notes

### Competing Interest Statement

The authors have declared no competing interest.

### Funding Statement

(DST-SERB) (EMR/2016/004829).

### Author Declarations

Institutional Review Board (IRB)/ ILBS

